# Deep Learning Model of Diastolic Dysfunction Risk Stratifies the Progression of Early-Stage Aortic Stenosis

**DOI:** 10.1101/2024.05.30.24308192

**Authors:** Márton Tokodi, Rohan Shah, Ankush Jamthikar, Neil Craig, Yasmin Hamirani, Grace Casaclang-Verzosa, Rebecca T. Hahn, Marc R. Dweck, Philippe Pibarot, Naveena Yanamala, Partho P. Sengupta

## Abstract

**Background:** The development and progression of aortic stenosis (AS) from aortic valve (AV) sclerosis is highly variable and difficult to predict.

**Objectives:** We investigated whether a previously validated echocardiography-based deep learning (DL) model assessing diastolic dysfunction (DD) could identify the latent risk associated with the development and progression of AS.

**Methods:** We evaluated 898 participants with AV sclerosis from the Atherosclerosis Risk in Communities (ARIC) cohort study and associated the DL-predicted probability of DD with two endpoints: (1) the new diagnosis of AS and (2) the composite of subsequent mortality or AV interventions. We performed validation in two additional cohorts: 1) patients with mild-to-moderate AS undergoing cardiac magnetic resonance (CMR) imaging and serial echocardiographic assessments (n=50), and (2) patients with AV sclerosis undergoing ^18^F-sodium fluoride (^18^F-NaF) and ^18^F-fluorodeoxyglucose positron emission tomography (PET) combined with computed tomography (CT) to assess valvular inflammation and calcification (n=18).

**Results:** In the ARIC cohort, a higher DL-predicted probability of DD was associated with the development of AS (adjusted HR: 3.482 [2.061 – 5.884], p<0.001) and subsequent mortality or AV interventions (adjusted HR: 7.033 [3.036 – 16.290], p<0.001). The multivariable Cox model (incorporating the DL-predicted probability of DD) derived from the ARIC cohort efficiently predicted the progression of AS (C-index: 0.798 [0.648 – 0.948]) in the CMR cohort. Moreover, the predictions of this multivariable Cox model correlated positively with valvular ^18^F-NaF mean standardized uptake values in the PET/CT cohort (r=0.62, p=0.008).

**Conclusions:** Assessment of DD using DL can stratify the latent risk associated with the progression of early-stage AS.

**CONDENSED ABSTRACT:** We investigated whether DD assessed using DL can predict the progression of early-stage AS. In 898 patients with AV sclerosis, the DL-predicted probability of DD was associated with the development of AS. The multivariable Cox model derived from these patients also predicted the progression of AS in an external cohort of patients with mild-to-moderate AS (n=50). Moreover, the predictions of this model correlated positively with PET/CT-derived valvular ^18^F-NaF uptake in an additional cohort of patients with AV sclerosis (n=18). These findings suggest that assessing DD using DL can stratify the latent risk associated with the progression of early-stage AS.

---

Aortic valve (AV) sclerosis, defined as the calcification and thickening of the AV that does not cause significant obstruction in transvalvular flow, is a common finding, and its prevalence increases from 25% at the age of 65 to 50% at 80 (1,2). AV sclerosis progresses to aortic stenosis (AS) in 10-15% of patients over 5 years (3–5) and is associated with adverse outcomes, such as coronary events and cardiovascular as well as all-cause mortality (1,6). Accordingly, the American College of Cardiology (ACC) and the American Heart Association (AHA) have recommended defining AS during its early stages as AV sclerosis (Stage A) and mild-to-moderate AS (Stage B) (7).

Several factors, including age, hyperlipidemia, diabetes, smoking, and hypertension, influence the progression of AV sclerosis and other myocardial processes like left ventricular (LV) remodeling and systolic and diastolic dysfunction (DD) (2,8–10). Notably, among different myocardial changes, the alterations in diastolic function – an active, adenosine triphosphate-dependent process – occur early and may explain the association of subclinical DD with AV sclerosis, independent of the aforementioned cardiovascular risk factors (11). Additionally, the compromised myocardial energetics and mitochondrial dysfunction associated with AS also contribute to DD (12–14).

Given these complex associations, machine learning may aid in identifying subsets of patients with AV sclerosis who face an elevated risk of progressing to AS. Nonetheless, previous machine learning and deep learning (DL) studies have primarily focused on the diagnosis (15–17), prognostication (18), and follow-up of patients with AS (19). Saliency maps in recent electrocardiogram (ECG)-based DL models highlighted a strong dependence on the diastolic phase, specifically the end of the T and U waves, for detecting AS, along with the future risk of developing AS in those with no AS at baseline (20–23). Moreover, the probability of AS predicted by these ECG-based models correlated positively with several echocardiographic features that assess the overall severity of DD (21). Similarly, saliency maps generated from a recent echocardiography-based DL model predicting the presence of AS from two-dimensional parasternal long-axis videos have demonstrated that the model also focused on other cardiac structures beyond the AV, and the model’s predictions correlated with echocardiographic markers of elevated diastolic filling pressures, left atrial dilation, and elevated pulmonary pressures (24).

Accordingly, we hypothesized that a DL model that integrates multiple echocardiographic features for assessing DD could identify the latent risk associated with the progression of early-stage AS. First, we investigated the associations between DD assessed using a previously validated DL model and the development of AS in patients with AV sclerosis (Stage A) from a population-based cohort. Then, we also confirmed the relevance of the DL-predicted DD probabilities in predicting the progression of AS in a second cohort of patients with mild-to-moderate AS (Stage B). Last, to gain insights into the underlying biological pathways, we also correlated the DL-predicted DD probabilities with the extent of myocardial fibrosis quantified using cardiac magnetic resonance (CMR) imaging in the second cohort and, in a third cohort of patients, we also assessed valvular calcification and inflammation using ^18^F-sodium fluoride (^18^F-NaF) and ^18^F-fluorodeoxyglucose (^18^F-FDG) positron emission tomography (PET) combined with computed tomography (CT).

## METHODS

### Population-based cohort – the ARIC cohort

We investigated the development of AS from AV sclerosis (Stage A) using data collected at visit 5 in the Atherosclerosis Risk in Communities (ARIC) cohort study (25). We analyzed the data of those who underwent an echocardiographic examination at visit 5 between 2011 and 2013 and whose data was available in the Biologic Specimen and Data Repository Information Coordinating Center (BioLINCC) database (Figure 1). From the 5,576 participants who fulfilled these criteria, we excluded those with congenital heart disease (n=20), hypertrophic obstructive cardiomyopathy (n=1), previously diagnosed AS (n=193), previous AV interventions (n=46), a prosthetic valve at any position (n=47), moderate or severe aortic regurgitation (n=24), or moderate or severe mitral regurgitation (n=108). In addition, we also excluded participants with missing values in key echocardiographic variables (n= 282) or no follow-up data (n=3).

**Figure 1.**
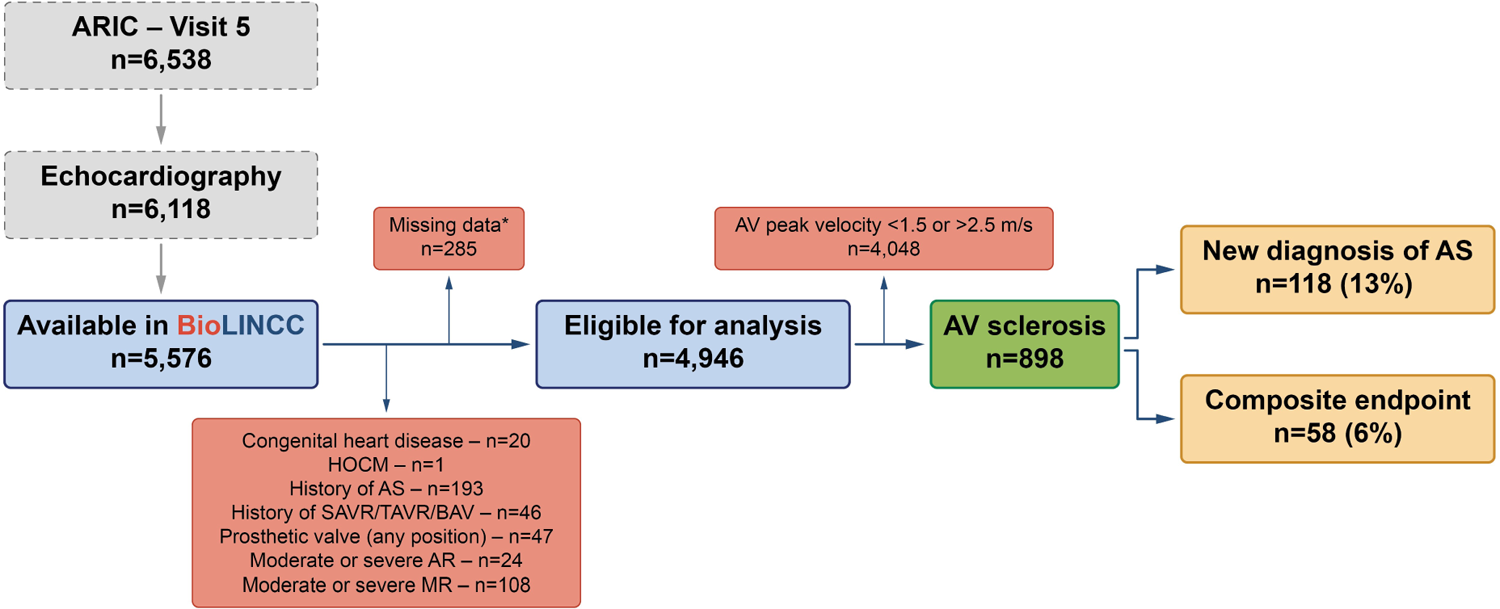
Patient selection flowchart of the ARIC cohort *Missing values in key echocardiographic variables or no follow-up data. Of the 4,946 participants eligible for analysis, 3,987 (81%) had an AV peak velocity lower than 1.5 m/s, and 61 (1%) had an AV peak velocity higher than 2.5 m/s at visit 5. AR – aortic regurgitation, ARIC – Atherosclerosis Risk in Communities study, AS – aortic stenosis, AV – aortic valve, BAV – bicuspid aortic valve, BioLINCC – Biologic Specimen and Data Repository Information Coordinating Center, HOCM – hypertrophic obstructive cardiomyopathy, MR – mitral regurgitation, SAVR – surgical aortic valve replacement, TAVR – transcatheter aortic valve replacement

### Echocardiographic protocol in the ARIC cohort

The echocardiographic protocol of visit 5 has been published previously (26) and is described briefly in the Supplemental Methods. AV sclerosis was defined as an AV peak velocity between 1.5 and 2.5 m/s (27). The presence and grade of DD were assessed based on the recommendations of the American Society of Echocardiography (ASE) and the European Association of Cardiovascular Imaging (EACVI) (28).

### DL-based assessment of DD

We used our previously published and thoroughly validated DL model that assesses DD based on nine routinely measured echocardiographic parameters: LV ejection fraction [LVEF], LV mass index [LVMi], left atrial volume index [LAVi], early mitral inflow velocity [E], late mitral inflow velocity (A), E/A, early diastolic mitral annular velocity at septal position [e’], E/e’, and tricuspid regurgitation peak velocity (TRV) (29,30). The output of the DL model is a single numeric value for each subject, denoting the probability of DD. Further details on the DL model and its external validation are provided in the Supplemental Methods and have been previously published (30). The model is publicly available online (31).

### Outcomes of interest in the ARIC cohort

The primary endpoint of our study was the new diagnosis of AS after visit 5, whereas the secondary endpoint was the composite of AV interventions and all-cause death after newly diagnosed AS. The time to event was measured from the date of the echocardiographic examination at visit 5. The new diagnosis of AS was established based on echocardiographic data available from abstracted hospitalizations or was defined as an event (hospitalization or death) with an International Classification of Diseases, 9th Revision (ICD-9) code of 424.1 or an ICD, 10th Revision (ICD-10) code of I35.0 or I35.2 in any position (Supplemental Table 1) (32). AV interventions were identified as hospitalizations with an ICD-9 procedure code of 35.01, 35.05, 35.06, 35.11, 35.21, 35.22, or 35.96 or an ICD-10 procedure code of 027Fx, 02NFx, 02QFx, or 02RFx in any position (Supplemental Table 2) (32). Participants not reaching these endpoints were followed up through December 31, 2018, date of death, or loss to follow-up, whichever occurred first.

### Multimodality external validation cohorts

#### CMR cohort

We applied the DL model to the participants of the “The Role of Myocardial Fibrosis in Patients With Aortic Stenosis” prospective observational study (NCT01755936) (33). All subjects underwent detailed clinical evaluation at baseline, including echocardiography and CMR imaging. Focal replacement fibrosis was assessed in all patients using late gadolinium enhancement (LGE), and diffuse fibrosis associated with extracellular volume expansion was assessed using myocardial T1 mapping. The details of the imaging protocol, image analysis, and validation against myocardial biopsy-derived histological myocardial fibrosis have been described previously (33). Patients were followed with annual echocardiographic examinations for two consecutive years. Progression of AS was defined as a worsening in the severity of AS (i.e., development of moderate or severe AS from mild or moderate AS, respectively). The severity of AS was determined based on AV mean gradient, AV peak velocity, AV area, and the dimensionless index.

#### PET/CT cohort

We also applied our DL model to the participants with AV sclerosis from the “Role of Active Valvular Calcification and Inflammation in Patients With Aortic Stenosis” observational cohort study (NCT01358513) (34). Participants in this study underwent echocardiography, non-contrast CT, and ^18^F-NaF and ^18^F-FDG PET/CT at baseline, with clinical and echocardiographic follow-up. The imaging protocols and image analysis techniques have been described previously (34). Twenty participants in this cohort had AV sclerosis (defined as an AV peak velocity between 1.5 and 2.5 m/s), but two participants were excluded from the current analysis due to missing echocardiographic data at baseline. One participant did not complete the ^18^F-NaF uptake analysis, while another did not undergo ^18^F-FDG uptake assessment.

### Statistical analysis

Continuous variables are expressed as median (interquartile range) or mean ± standard deviation, while categorical variables are reported as frequencies and percentages. The normality of continuous variables was checked using the Shapiro-Wilk test. The characteristics of patient subgroups were compared using unpaired Student’s t-test or Mann-Whitney U test for continuous variables and Chi-squared or Fisher’s exact test for categorical variables, as appropriate. The event-free survival of subgroups was visualized on Kaplan-Meier curves, and Log-rank tests were performed for comparison. Univariable and multivariable Cox proportional hazards models were used to compute hazard ratios (HRs) with 95% confidence intervals (CIs). To better understand the effects that the DL-predicted probability has on the primary endpoint, we computed pointwise estimates of the univariable and multivariable HR curves and the corresponding confidence limits using the smoothHR R package (35). The optimal degree of freedom was obtained by minimizing the corrected Akaike information criterion. The probability value where the lower bound of the confidence band of the multivariable HR intersects the HR of 1 was used as the cut-off value to discriminate between patients with high and low risk of developing AS. To demonstrate the incremental prognostic value of the DL-derived predictions over other conventional covariates, sequential (i.e., nested) Cox regression models were also constructed, which were then compared using the likelihood ratio test, Harrell’s C-index (i.e., concordance index), integrated discrimination index (IDI), and net reclassification index (NRI). The final multivariable Cox regression model built using the ARIC dataset to predict the development of AS was applied to the external validation cohorts. We calculated the linear predictor as the weighted sum of the mean-centered independent variables (i.e., predictors) in the Cox regression model, where the weights were the regression coefficients (Supplemental Methods). Pearson’s correlation coefficients were calculated to assess the strength of linear associations between the linear predictor and the ^18^F-NaF and ^18^F-FDG maximum and mean standardized uptake values (SUV).

A P-value of <0.05 was considered statistically significant. All statistical analyses were performed in R (version 4.1.2, R Foundation for Statistical Computing, Vienna, Austria). The utilized versions of the R packages are documented in Supplemental Table 3.

### Ethical approval

All participants of the ARIC cohort study provided written informed consent, and the institutional review boards associated with each field center approved the study protocol. The two studies used for external validation (NCT01755936 and NCT01358513) were approved by the corresponding local research ethics committees, and written informed consent was obtained from all patients. The protocol of the current analysis conforms with the principles outlined in the Declaration of Helsinki, and it was approved by the Institutional Review Board of Rutgers Biomedical and Health Sciences (study identification number: Pro2021001505).

## RESULTS

### Clinical characteristics and outcomes of the participants in the ARIC cohort

Among the 5,576 participants (75 [71 – 79] years, 57% female, 19% black), 898 (16%) had AV sclerosis (Figure 1, Table 1). Over the median follow-up duration of 6.4 (5.7 – 6.9) years, 118 of 898 AV sclerosis patients (13%) developed AS, whereas 58 (6%) reached the composite endpoint. Of the 118 new diagnoses of AS, 72 (61%) were established using echocardiography, while the remaining were identified based on ICD codes.

**Table 1.** Clinical and echocardiographic characteristics of patients with high and low risk of progressing to AS in the ARIC cohort.

|  | Missing<br>n (%) | All participants<br>with AV sclerosis<br>n=898 | High-risk group<br>n=346 | Low-risk group<br>n=552 | P-value |
| --- | --- | --- | --- | --- | --- |
| **Demographics, vitals, risk factors** |  |  |  |  |  |
| Age, years | 0 (0) | 76 (72 – 80) | 77 (73 – 82) | 75 (71 – 79) | <0.001 |
| Male sex | 0 (0) | 404 (45) | 174 (50) | 230 (42) | 0.014 |
| Black race | 0 (0) | 152 (17) | 71 (21) | 81 (15) | 0.029 |
| BMI, kg/m<sup>2</sup> | 3 (0) | 28.9 (25.9 – 33.1) | 29.9 (26.3 – 34.4) | 28.5 (25.7 – 32.7) | 0.003 |
| BSA, m<sup>2</sup> | 0 (0) | 1.89 (1.74 – 2.04) | 1.93 (1.77 – 2.08) | 1.88 (1.73 – 2.03) | 0.020 |
| SBP, mmHg | 2 (0) | 130 (119 – 142) | 133 (119 – 146) | 128 (119 – 140) | 0.009 |
| DBP, mmHg | 2 (0) | 65 (58 – 72) | 65 (57 – 73) | 65 (58 – 71) | 0.812 |
| HR, 1/min | 2 (0) | 63 (57 – 71) | 62 (55 – 70) | 64 (58 – 71) | 0.007 |
| Hypertension | 0 (0) | 769 (86) | 315 (91) | 454 (82) | <0.001 |
| Diabetes | 0 (0) | 353 (39) | 156 (45) | 197 (36) | 0.006 |
| Chronic kidney disease | 0 (0) | 283 (32) | 135 (39) | 148 (27) | <0.001 |
| Smoking status | 66 (7) |  |  |  |  |
| Never smoker |  | 335 (40) | 122 (39) | 213 (41) |  |
| Current smoker |  | 47 (6) | 17 (5) | 30 (6) |  |
| Former smoker |  | 450 (54) | 177 (56) | 273 (53) | 0.695 |
| History of HF | 0 (0) | 163 (18) | 103 (30) | 60 (11) | <0.001 |
| History of CHD | 0 (0) | 179 (20) | 101 (29) | 78 (14) | <0.001 |
| History of AF | 0 (0) | 95 (11) | 61 (18) | 34 (6) | <0.001 |
| History of stroke | 0 (0) | 60 (7) | 33 (10) | 27 (5) | 0.010 |
| **Medications** |  |  |  |  |  |
| Antihypertensive medications | 0 (0) | 718 (80) | 299 (86) | 419 (76) | <0.001 |
| Antidiabetic medications | 3 (0) | 195 (22) | 97 (28) | 98 (18) | <0.001 |
| Statin | 6 (1) | 512 (57) | 203 (59) | 309 (56) | 0.434 |
| Cholesterol-lowering medications | 6 (1) | 558 (63) | 221 (64) | 337 (61) | 0.399 |
| **Laboratory results** |  |  |  |  |  |
| Creatinine, mg/dL | 5 (1) | 0.93 (0.79 – 1.12) | 0.98 (0.82 – 1.19) | 0.91 (0.77 – 1.07) | <0.001 |
| GFR, mL/min/1.73m<sup>2</sup> | 5 (1) | 70 (58 – 82) | 68 (54 – 81) | 70 (60 – 84) | 0.001 |
| Uric acid, mg/dL | 5 (1) | 5.8 (4.9 – 6.7) | 5.9 (4.9 – 7.0) | 5.7 (4.8 – 6.6) | 0.095 |
| Fasting glucose, mg/dL | 11 (1) | 107 (98 – 122) | 109 (98 – 125) | 106 (98 – 120) | 0.128 |
| Hemoglobin A1C, % | 7 (1) | 5.7 (5.5 – 6.1) | 5.8 (5.5 – 6.4) | 5.7 (5.5 – 6.0) | 0.006 |
| Triglyceride, mg/dL | 6 (1) | 112 (85 – 156) | 109 (83 – 148) | 113 (87 – 158) | 0.128 |
| Total chol., mg/dL | 6 (1) | 174 (150 – 201) | 166 (136 – 197) | 179 (155 – 204) | <0.001 |
| LDL-C, mg/dL | 12 (1) | 98 (77 – 122) | 93 (69 – 121) | 102 (81 – 123) | <0.001 |
| HDL-C, mg/dL | 6 (1) | 50 (41 – 58) | 48 (39 – 56) | 51 (42 – 58) | <0.001 |
| NT-proBNP, pg/mL | 29 (3) | 143 (75 – 290) | 240 (114 – 478) | 113 (65 – 205) | <0.001 |
| hs-Troponin T, ng/mL | 5 (1) | 0.01 (0.01 – 0.02) | 0.01 (0.01 – 0.02) | 0.01 (0.01 – 0.01) | <0.001 |
| hs-CRP, mg/dL | 6 (1) | 2.17 (1.03 – 4.60) | 2.33 (1.20 – 4.87) | 2.07 (1.01 – 4.35) | 0.123 |
| Hemoglobin, g/dL | 27 (3) | 13.3 (12.4 – 14.3) | 13.3 (12.4 – 14.3) | 13.4 (12.5 – 14.3) | 0.524 |
| RDW, % | 27 (3) | 14.2 (13.6 – 15.0) | 14.3 (13.6 – 15.0) | 14.2 (13.6 – 14.9) | 0.124 |
| **Echocardiographic parameters** |  |  |  |  |  |
| IVSd, cm | 0 (0) | 1.05 (0.95 – 1.18) | 1.15 (1.02 – 1.26) | 1.00 (0.93 – 1.12) | <0.001 |
| LVPWd, cm | 0 (0) | 0.94 (0.87 – 1.03) | 1.01 (0.92 – 1.13) | 0.90 (0.85 – 0.97) | <0.001 |
| LVIDd, cm | 0 (0) | 4.48 ± 0.53 | 4.70 ± 0.56 | 4.35 ± 0.47 | <0.001 |
| LVIDs, cm | 0 (0) | 2.60 (2.25 – 2.95) | 2.79 (2.38 – 3.19) | 2.51 (2.17 – 2.80) | <0.001 |
| LVMi, g/m<sup>2</sup> | 0 (0) | 79.9 (69.2 – 94.9) | 95.5 (81.5 – 110.) | 73.3 (65.6 – 83.0) | <0.001 |
| LVEDVi, mL/m<sup>2</sup> | 0 (0) | 44.1 (37.5 – 51.3) | 48.43 (41.3 – 56.1) | 41.58 (36.1 – 48.6) | <0.001 |
| LVESVi, mL/m<sup>2</sup> | 0 (0) | 14.4 (11.5 – 18.1) | 17.2 (13.9 – 21.3) | 13.2 (10.8 – 15.9) | <0.001 |
| LVEF, % | 0 (0) | 67.1 (62.7 – 70.8) | 63.9 (60.0 – 68.3) | 68.3 (65.4 – 71.7) | <0.001 |
| LVGLS, % | 30 (3) | -18.6<br>(-20.1 to -16.8) | -17.5<br>(-19.2 to -15.7) | -19.1<br>(-20.4 to -17.7) | <0.001 |
| LAVi, mL/m<sup>2</sup> | 0 (0) | 26.9 (22.4 – 32.5) | 31.1 (25.7 – 37.6) | 24.9 (21.0 – 29.7) | <0.001 |

**Table 1 Continued**
|  | Missing<br>n (%) | All participants<br>with AV sclerosis<br>n=898 | Participants with<br>events<br>n=346 | Participants<br>without events<br>n=552 | P-value |
| --- | --- | --- | --- | --- | --- |
| AV peak velocity, m/s | 0 (0) | 1.66 (1.56 – 1.86) | 1.67 (1.55 – 1.89) | 1.65 (1.56 – 1.84) | 0.245 |
| AV mean gradient,<br>mmHg | 0 (0) | 5.2 (4.5 – 6.5) | 5.4 (4.6 – 6.7) | 5.2 (4.5 – 6.3) | 0.023 |
| Dimensionless index | 0 (0) | 0.68 (0.57 – 0.78) | 0.64 (0.52 – 0.74) | 0.70 (0.59 – 0.81) | <0.001 |
| E, cm/s | 0 (0) | 71 (60 – 83) | 75 (61 – 89) | 70 (59 – 80) | <0.001 |
| E/A | 35 (4) | 0.8 (0.7 – 1.0) | 0.8 (0.6 – 0.9) | 0.8 (0.7 – 1.0) | 0.028 |
| e' (septal), cm/s | 0 (0) | 5.6 (4.8 – 6.6) | 4.8 (4.1 – 5.3) | 6.2 (5.3 – 7.2) | <0.001 |
| E/e' (septal) | 0 (0) | 12.4 (10.2 – 15.6) | 15.5 (12.7 – 19.4) | 11.04 (9.5 – 13.1) | <0.001 |
| TRV, m/s | 334 (37) | 2.45 (2.25 – 2.65) | 2.49 (2.25 – 2.73) | 2.42 (2.25 – 2.60) | 0.004 |
| Guideline-based DD | 0 (0) |  |  |  |  |
| No DD |  | 222 (25) | 30 (9) | 192 (35) |  |
| Grade I |  | 459 (51) | 179 (52) | 280 (51) |  |
| Grade II |  | 78 (9) | 64 (18) | 14 (3) |  |
| Grade III |  | 6 (1) | 6 (2) | 0 (0) |  |
| Indeterminate |  | 133 (15) | 67 (19) | 66 (12) | <0.001 |
| **DL-predicted**<br>**probability of DD** | 0 (0) | 0.50 (0.04 – 0.97) | 0.99 (0.94 – 1.00) | 0.07 (0.01 – 0.38) | <0.001 |
| **DL-predicted high risk** | 0 (0) | 346 (39) | 346 (100) | 0 (0) | <0.001 |

Continuous variables are presented as median (interquartile range) and categorical variables as n (%). Comparisons between the high-risk and low-risk phenogroups were performed using unpaired Student's t-test or Mann-Whitney U test for continuous variables, Chi-squared or Fisher's exact test for categorical variables, as appropriate.
A – late mitral inflow velocity, AF – atrial fibrillation, ARIC – Atherosclerosis Risk in Communities study, AS – aortic stenosis, AV – aortic valve, BMI – body mass index, BSA – body surface area, CHD – coronary heart disease, CRP – C-reactive protein, DBP – diastolic blood pressure, DD – diastolic dysfunction, DL – deep learning, E – early mitral inflow velocity, e' – early diastolic mitral annular velocity, GFR – glomerular filtration rate, HDL-C – high-density lipoprotein cholesterol, HF – heart failure, HR – heart rate, hs – high-sensitivity, IVSd – thickness of the interventricular septum at end-diastole, LAVi – left atrial volume index, LDL-C – low-density lipoprotein cholesterol, LVEDVi – left ventricular end-diastolic volume index, LVEF – left ventricular ejection fraction, LVESVi – left ventricular end-systolic volume index, LVIDd – left ventricular internal diameter at end-diastole, LVIDs – left ventricular internal diameter at end-systole, LVGLS – left ventricular global longitudinal strain, LVMI – left ventricular mass index, LVPWd – thickness of the left ventricular posterior wall at end-diastole, NT-proBNP – N-terminal pro-brain natriuretic peptide, RDW – red cell distribution width, SBP – systolic blood pressure, TRV – tricuspid regurgitation peak velocity

### Associations of the DL-derived predictions with outcomes in the ARIC cohort

The univariable and multivariable HR curves revealed that using the DL-predicted probability value of 0 as the reference, the risk of developing AS became significant at the probability value of 0.84 and 0.81, respectively, and continuously increased until the probability value of 1 (Figure 2). Using the latter as a cut-off, we assigned each subject either to the low-risk (probability of <0.81) or the high-risk group (probability of ≥0.81) (Table 1).

**Figure 2.**
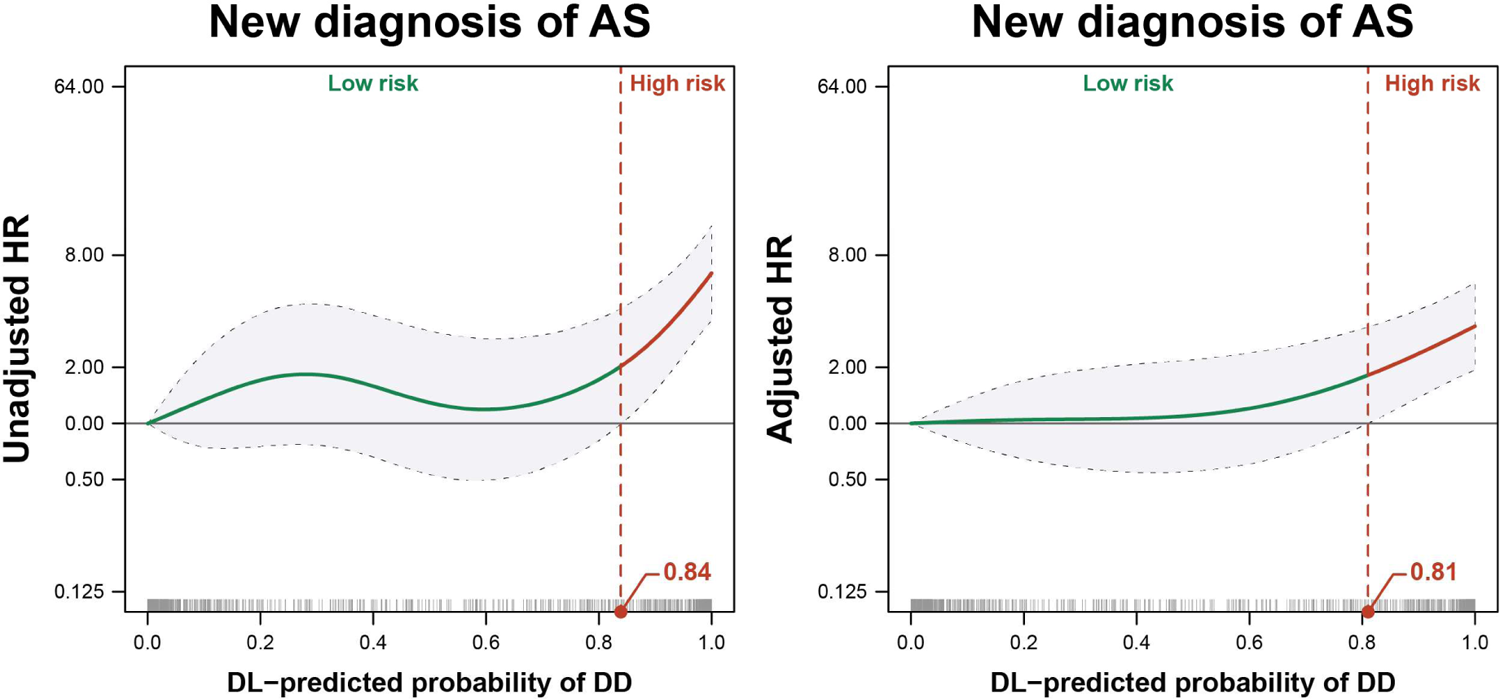
The dependence of the risk of developing AS on the DL-predicted probability of DD in the ARIC cohort The probability value of 0 was taken as the reference. The red vertical dashed line indicates the probability value where the lower bound of the confidence band intersects the HR of 1. DD – diastolic dysfunction, DL – deep learning, HR – hazard ratio; other abbreviations as in Figure 1.

When we plotted and compared the event-free survival of the two groups, we observed that a higher proportion of high-risk compared to low-risk patients was diagnosed with AS and reached the composite endpoint during follow-up (Figure 3A). We could also observe a significant separation between the survival curves of the two risk groups when we used the default probability threshold of 0.5 (Figure 3B). Moreover, we also demonstrated in univariable Cox regression analysis that the DL-predicted probability is a significant predictor of both outcomes of interest (Tables 2 and 3). Using a multivariable Cox regression model including clinical variables and AV peak velocity as covariates, we also confirmed that the DL-predicted probability was an independent predictor of the new diagnosis of AS (Model AS3, Table 4). In addition, the probability values were also associated with this outcome of interest in multivariable models that included several laboratory parameters or smoking status, chronic kidney disease, and medications besides AV peak velocity (Supplemental Tables 4 and 5). Although we could only include age, sex, race, and AV peak velocity as a covariate due to the limited number of participants reaching the composite endpoint, the DL-predicted probability was still found to be an independent predictor of this endpoint as well (Supplemental Table 6). We also performed a sensitivity analysis by applying right censoring if the new diagnosis of AS was not ascertained by echocardiography and found that the DL-predicted probability of DD was still associated with the new diagnosis of AS (Supplemental Table 7). The incremental value of DL-predicted DD probability over ASE/EACVI guideline-based DD grading is presented in the Supplementary Appendix (Supplemental Results, Supplemental Table 8, and Supplemental Figures 1 and 2).

**Figure 3.**
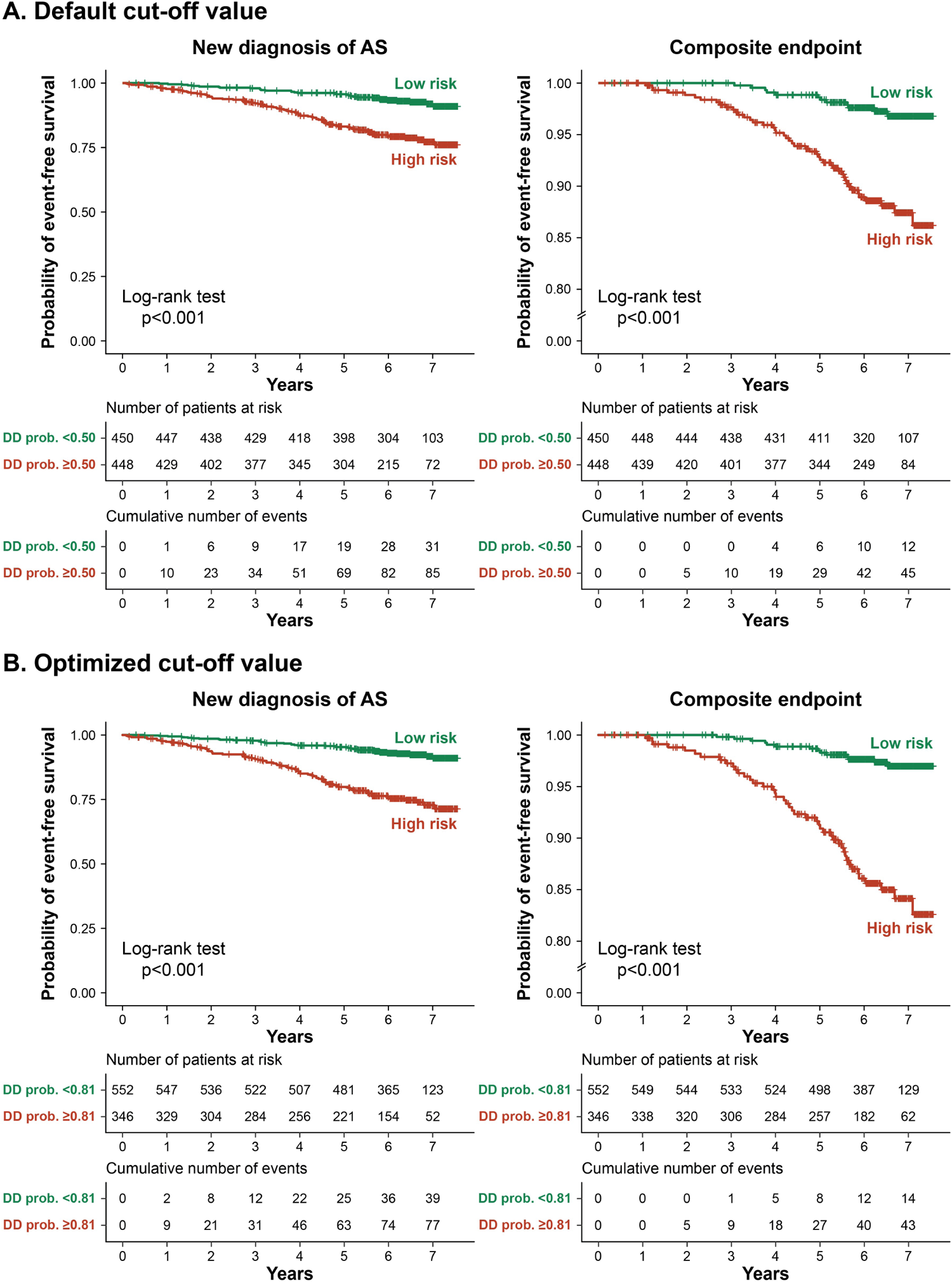
Kaplan-Meier curves showing the event-free survival of patients with AV sclerosis in the ARIC cohort In panel A, the default cut-off value (i.e., the DD probability value of 0.50) was used to stratify patients into low-risk and high-risk groups. In panel B, the cut-off value optimized in the ARIC cohort (i.e., the DD probability value of 0.81) was used to stratify patients into low-risk and high-risk groups. Abbreviations as in Figure 1.

**Table 2.**
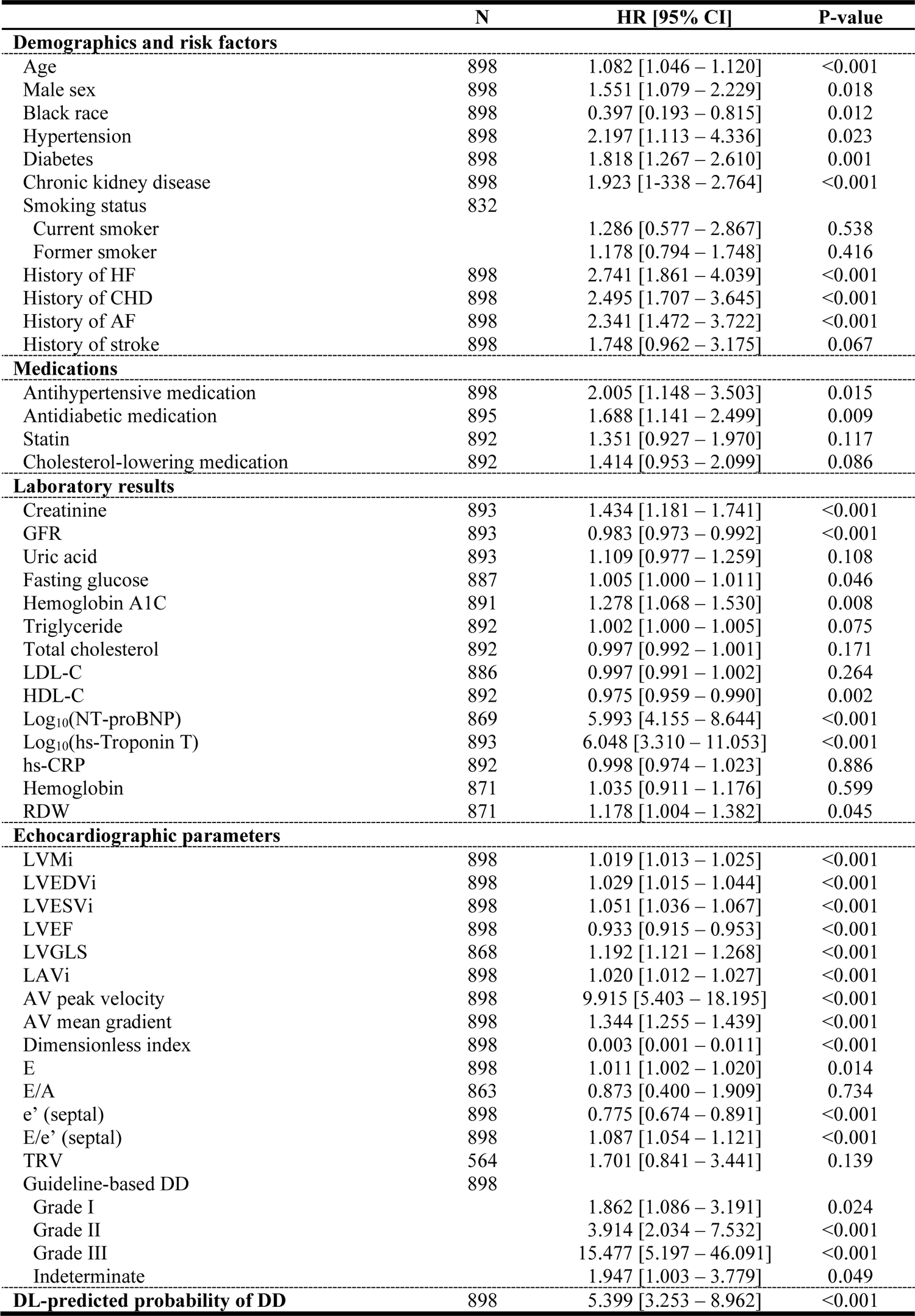
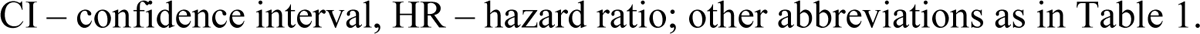
Univariable Cox regression models for predicting the new diagnosis of AS in patients with AV sclerosis in the ARIC cohort.

|  | N | HR [95% CI] | P-value |
| --- | --- | --- | --- |
| **Demographics and risk factors** |  |  |  |
| Age | 898 | 1.082 [1.046 – 1.120] | <0.001 |
| Male sex | 898 | 1.551 [1.079 – 2.229] | 0.018 |
| Black race | 898 | 0.397 [0.193 – 0.815] | 0.012 |
| Hypertension | 898 | 2.197 [1.113 – 4.336] | 0.023 |
| Diabetes | 898 | 1.818 [1.267 – 2.610] | 0.001 |
| Chronic kidney disease | 898 | 1.923 [1.338 – 2.764] | <0.001 |
| Smoking status | 832 |  |  |
| Current smoker |  | 1.286 [0.577 – 2.867] | 0.538 |
| Former smoker |  | 1.178 [0.794 – 1.748] | 0.416 |
| History of HF | 898 | 2.741 [1.861 – 4.039] | <0.001 |
| History of CHD | 898 | 2.495 [1.707 – 3.645] | <0.001 |
| History of AF | 898 | 2.341 [1.472 – 3.722] | <0.001 |
| History of stroke | 898 | 1.748 [0.962 – 3.175] | 0.067 |
| **Medications** |  |  |  |
| Antihypertensive medication | 898 | 2.005 [1.148 – 3.503] | 0.015 |
| Antidiabetic medication | 895 | 1.688 [1.141 – 2.499] | 0.009 |
| Statin | 892 | 1.351 [0.927 – 1.970] | 0.117 |
| Cholesterol-lowering medication | 892 | 1.414 [0.953 – 2.099] | 0.086 |
| **Laboratory results** |  |  |  |
| Creatinine | 893 | 1.434 [1.181 – 1.741] | <0.001 |
| GFR | 893 | 0.983 [0.973 – 0.992] | <0.001 |
| Uric acid | 893 | 1.109 [0.977 – 1.259] | 0.108 |
| Fasting glucose | 887 | 1.005 [1.000 – 1.011] | 0.046 |
| Hemoglobin A1C | 891 | 1.278 [1.068 – 1.530] | 0.008 |
| Triglyceride | 892 | 1.002 [1.000 – 1.005] | 0.075 |
| Total cholesterol | 892 | 0.997 [0.992 – 1.001] | 0.171 |
| LDL-C | 886 | 0.997 [0.991 – 1.002] | 0.264 |
| HDL-C | 892 | 0.975 [0.959 – 0.990] | 0.002 |
| Log<sub>10</sub>(NT-proBNP) | 869 | 5.993 [4.155 – 8.644] | <0.001 |
| Log<sub>10</sub>(hs-Troponin T) | 893 | 6.048 [3.310 – 11.053] | <0.001 |
| hs-CRP | 892 | 0.998 [0.974 – 1.023] | 0.886 |
| Hemoglobin | 871 | 1.035 [0.911 – 1.176] | 0.599 |
| RDW | 871 | 1.178 [1.004 – 1.382] | 0.045 |
| **Echocardiographic parameters** |  |  |  |
| LVMi | 898 | 1.019 [1.013 – 1.025] | <0.001 |
| LVEDVi | 898 | 1.029 [1.015 – 1.044] | <0.001 |
| LVESVi | 898 | 1.051 [1.036 – 1.067] | <0.001 |
| LVEF | 898 | 0.933 [0.915 – 0.953] | <0.001 |
| LVGLS | 868 | 1.192 [1.121 – 1.268] | <0.001 |
| LAVi | 898 | 1.020 [1.012 – 1.027] | <0.001 |
| AV peak velocity | 898 | 9.915 [5.403 – 18.195] | <0.001 |
| AV mean gradient | 898 | 1.344 [1.255 – 1.439] | <0.001 |
| Dimensionless index | 898 | 0.003 [0.001 – 0.011] | <0.001 |
| E | 898 | 1.011 [1.002 – 1.020] | 0.014 |
| E/A | 863 | 0.873 [0.400 – 1.909] | 0.734 |
| e' (septal) | 898 | 0.775 [0.674 – 0.891] | <0.001 |
| E/e' (septal) | 898 | 1.087 [1.054 – 1.121] | <0.001 |
| TRV | 564 | 1.701 [0.841 – 3.441] | 0.139 |
| Guideline-based DD | 898 |  |  |
| Grade I |  | 1.862 [1.086 – 3.191] | 0.024 |
| Grade II |  | 3.914 [2.034 – 7.532] | <0.001 |
| Grade III |  | 15.477 [5.197 – 46.091] | <0.001 |
| Indeterminate |  | 1.947 [1.003 – 3.779] | 0.049 |
| **DL-predicted probability of DD** | 898 | 5.399 [3.253 – 8.962] | <0.001 |

CI – confidence interval, HR – hazard ratio; other abbreviations as in Table 1.

**Table 3.**
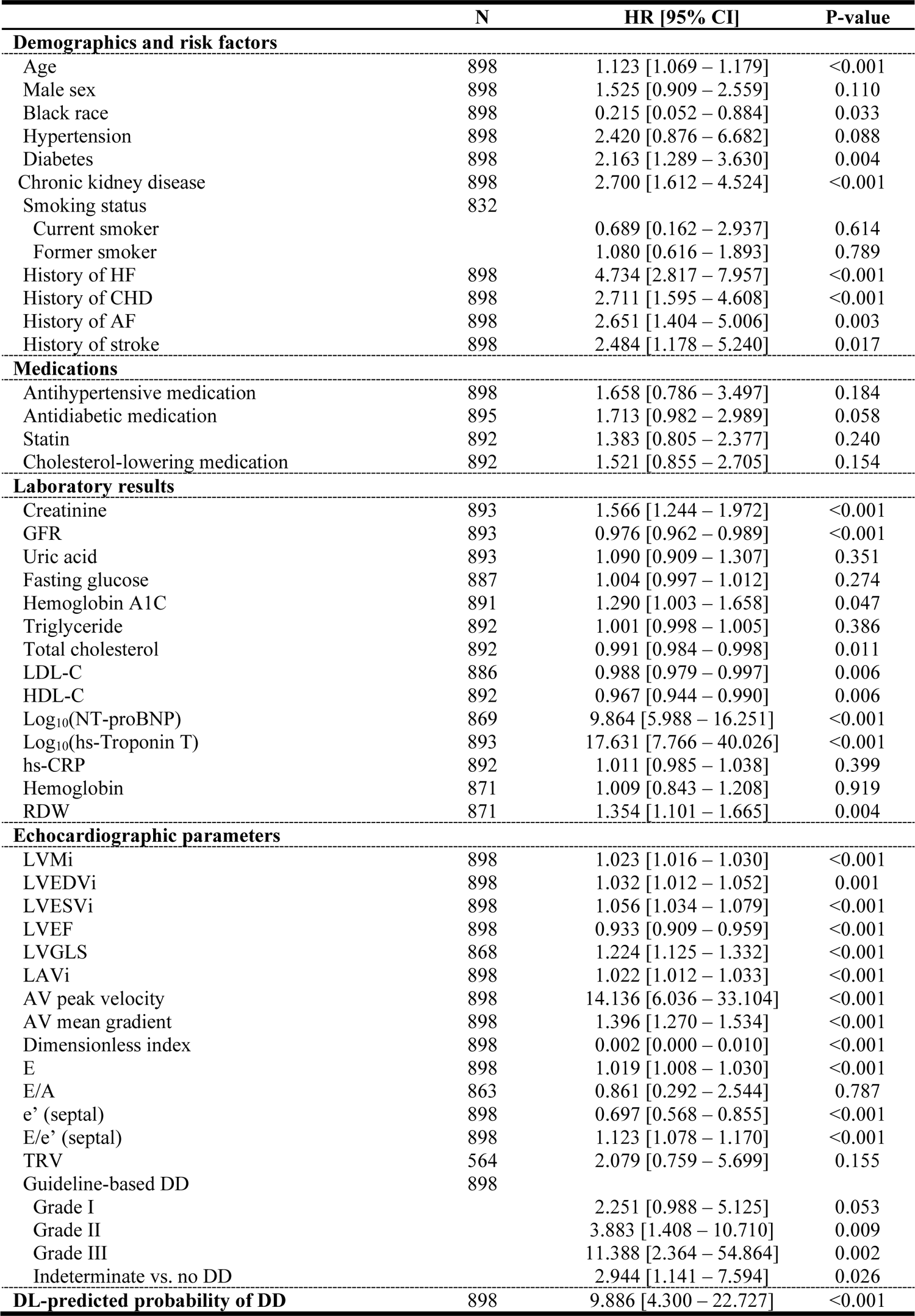
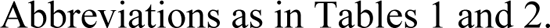
Univariable and multivariable Cox regression models for predicting the composite endpoint in patients with AV sclerosis in the ARIC cohort.

|  | N | HR [95% CI] | P-value |
| --- | --- | --- | --- |
| **Demographics and risk factors** |  |  |  |
| Age | 898 | 1.123 [1.069 – 1.179] | <0.001 |
| Male sex | 898 | 1.525 [0.909 – 2.559] | 0.110 |
| Black race | 898 | 0.215 [0.052 – 0.884] | 0.033 |
| Hypertension | 898 | 2.420 [0.876 – 6.682] | 0.088 |
| Diabetes | 898 | 2.163 [1.289 – 3.630] | 0.004 |
| Chronic kidney disease | 898 | 2.700 [1.612 – 4.524] | <0.001 |
| Smoking status | 832 |  |  |
| Current smoker |  | 0.689 [0.162 – 2.937] | 0.614 |
| Former smoker |  | 1.080 [0.616 – 1.893] | 0.789 |
| History of HF | 898 | 4.734 [2.817 – 7.957] | <0.001 |
| History of CHD | 898 | 2.711 [1.595 – 4.608] | <0.001 |
| History of AF | 898 | 2.651 [1.404 – 5.006] | 0.003 |
| History of stroke | 898 | 2.484 [1.178 – 5.240] | 0.017 |
| **Medications** |  |  |  |
| Antihypertensive medication | 898 | 1.658 [0.786 – 3.497] | 0.184 |
| Antidiabetic medication | 895 | 1.713 [0.982 – 2.989] | 0.058 |
| Statin | 892 | 1.383 [0.805 – 2.377] | 0.240 |
| Cholesterol-lowering medication | 892 | 1.521 [0.855 – 2.705] | 0.154 |
| **Laboratory results** |  |  |  |
| Creatinine | 893 | 1.566 [1.244 – 1.972] | <0.001 |
| GFR | 893 | 0.976 [0.962 – 0.989] | <0.001 |
| Uric acid | 893 | 1.090 [0.909 – 1.307] | 0.351 |
| Fasting glucose | 887 | 1.004 [0.997 – 1.012] | 0.274 |
| Hemoglobin A1C | 891 | 1.290 [1.003 – 1.658] | 0.047 |
| Triglyceride | 892 | 1.001 [0.998 – 1.005] | 0.386 |
| Total cholesterol | 892 | 0.991 [0.984 – 0.998] | 0.011 |
| LDL-C | 886 | 0.988 [0.979 – 0.997] | 0.006 |
| HDL-C | 892 | 0.967 [0.944 – 0.990] | 0.006 |
| Log<sub>10</sub>(NT-proBNP) | 869 | 9.864 [5.988 – 16.251] | <0.001 |
| Log<sub>10</sub>(hs-Troponin T) | 893 | 17.631 [7.766 – 40.026] | <0.001 |
| hs-CRP | 892 | 1.011 [0.985 – 1.038] | 0.399 |
| Hemoglobin | 871 | 1.009 [0.843 – 1.208] | 0.919 |
| RDW | 871 | 1.354 [1.101 – 1.665] | 0.004 |
| **Echocardiographic parameters** |  |  |  |
| LVMi | 898 | 1.023 [1.016 – 1.030] | <0.001 |
| LVEDVi | 898 | 1.032 [1.012 – 1.052] | 0.001 |
| LVESVi | 898 | 1.056 [1.034 – 1.079] | <0.001 |
| LVEF | 898 | 0.933 [0.909 – 0.959] | <0.001 |
| LVGLS | 868 | 1.224 [1.125 – 1.332] | <0.001 |
| LAVi | 898 | 1.022 [1.012 – 1.033] | <0.001 |
| AV peak velocity | 898 | 14.136 [6.036 – 33.104] | <0.001 |
| AV mean gradient | 898 | 1.396 [1.270 – 1.534] | <0.001 |
| Dimensionless index | 898 | 0.002 [0.000 – 0.010] | <0.001 |
| E | 898 | 1.019 [1.008 – 1.030] | <0.001 |
| E/A | 863 | 0.861 [0.292 – 2.544] | 0.787 |
| e' (septal) | 898 | 0.697 [0.568 – 0.855] | <0.001 |
| E/e' (septal) | 898 | 1.123 [1.078 – 1.170] | <0.001 |
| TRV | 564 | 2.079 [0.759 – 5.699] | 0.155 |
| Guideline-based DD | 898 |  |  |
| Grade I |  | 2.251 [0.988 – 5.125] | 0.053 |
| Grade II |  | 3.883 [1.408 – 10.710] | 0.009 |
| Grade III |  | 11.388 [2.364 – 54.864] | 0.002 |
| Indeterminate vs. no DD |  | 2.944 [1.141 – 7.594] | 0.026 |
| **DL-predicted probability of DD** | 898 | 9.886 [4.300 – 22.727] | <0.001 |

Abbreviations as in Tables 1 and 2.

**Table 4.** Sequential multivariable Cox regression models for predicting the new diagnosis of AS in patients with AV sclerosis in the ARIC cohort.

|  | **Model AS1**<br>(C-index: 0.709, AIC: 1,498) |  | **Model AS2**<br>(C-index: 0.747, AIC: 1,469) |  | **Model AS3**<br>(C-index: 0.773, AIC: 1,446) |  |
| --- | --- | --- | --- | --- | --- | --- |
|  | **HR [95% CI]** | **P-value** | **HR [95% CI]** | **P-value** | **HR [95% CI]** | **P-value** |
| Age, years | 1.061<br>[1.025 – 1.099] | <0.001 | 1.054<br>[1.019 – 1.091] | 0.002 | 1.043<br>[1.008 – 1.079] | 0.017 |
| Male sex | 1.179<br>[0.806 – 1.726] | 0.397 | 1.074<br>[0.728 – 1.584] | 0.720 | 1.007<br>[0.680 – 1.490] | 0.974 |
| Black race | 0.353<br>[0.169 – 0.738] | 0.006 | 0.383<br>[0.183 – 0.804] | 0.011 | 0.367<br>[0.174 – 0.772] | 0.008 |
| Hypertension | 1.781<br>[0.892 – 3.555] | 0.102 | 1.632<br>[0.817 – 3.260] | 0.165 | 1.471<br>[0.735 – 2.945] | 0.276 |
| Diabetes | 1.569<br>[1.085 – 2.268] | 0.017 | 1.555<br>[1.075 – 2.250] | 0.019 | 1.474<br>[1.016 – 2.139] | 0.041 |
| History of HF | 2.125<br>[1.373 – 3.289] | <0.001 | 2.045<br>[1.331 – 3.143] | 0.001 | 1.784<br>[1.160 – 2.742] | 0.008 |
| History of CHD | 1.427<br>[0.928 – 2.195] | 0.106 | 1.309<br>[0.850 – 2.016] | 0.221 | 1.210<br>[0.783 – 1.868] | 0.391 |
| History of AF | 1.474<br>[0.912 – 2.384] | 0.114 | 1.419<br>[0.881 – 2.283] | 0.150 | 1.328<br>[0.828 – 2.130] | 0.240 |
| AV peak velocity, m/s |  |  | 6.412<br>[3.427 – 11.998] | <0.001 | 5.689<br>[3.075 – 10.524] | <0.001 |
| DL-predicted probability of DD |  |  |  |  | 3.482<br>[2.061 – 5.884] | <0.001 |

AIC – Akaike information criterion; other abbreviations as in Tables 1 and 2.

### Incremental prognostic value of the DL-derived predictions over conventional risk factors

In predicting the new diagnosis of AS, the DL-predicted probability of DD showed incremental prognostic value over clinical variables and AV peak velocity with significant improvement in Harrel’s C-index, IDI, and NRI (Figure 4, Supplemental Table 9). It also had an incremental value over age, sex, race, and AV peak velocity for predicting the composite endpoint based on these three indices (Figure 4, Supplemental Table 10).

**Figure 4.**
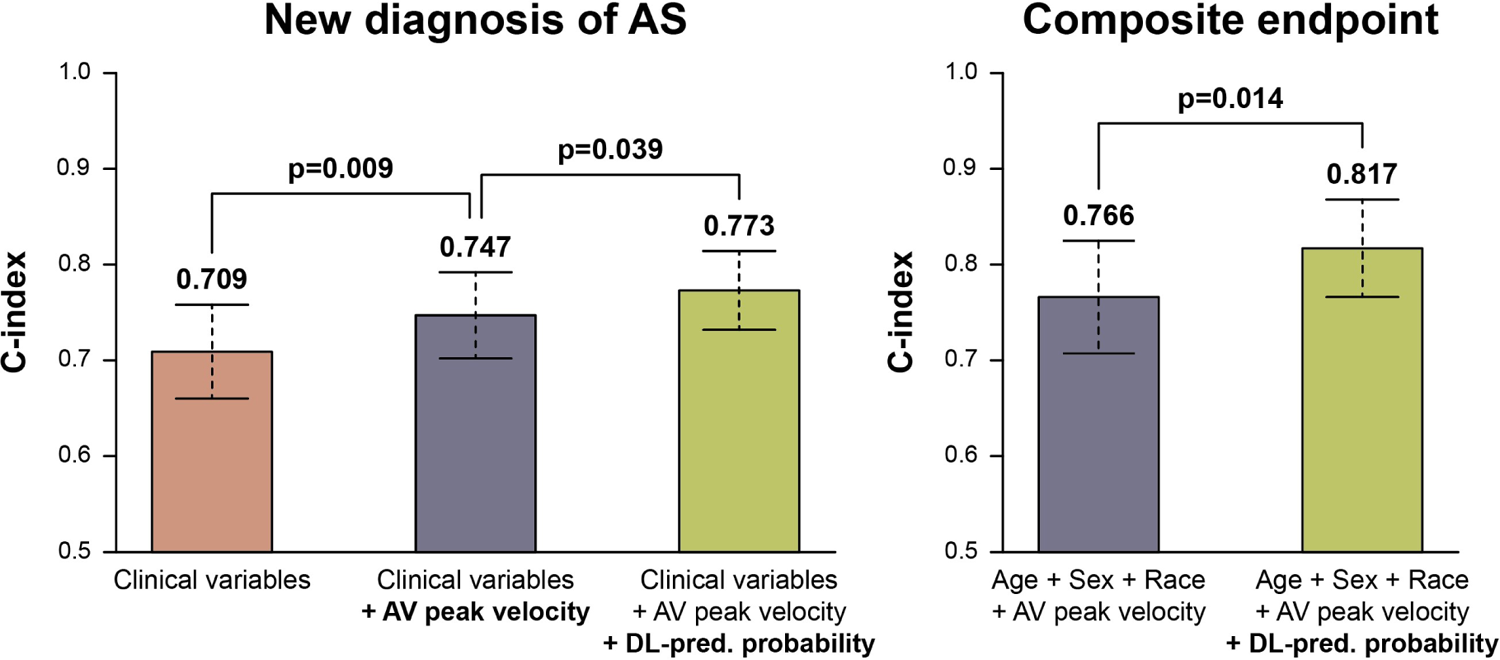
Sequential Cox regression models for predicting the new diagnosis of AS and the composite endpoint in patients with AV sclerosis in the ARIC cohort Clinical variables include age, sex, race, hypertension, diabetes, history of heart failure, history of coronary heart disease, and history of atrial fibrillation. Abbreviations as in Figures 1 and 2.

### External validation

#### CMR cohort

A total of 50 patients with Stage B AS (24 with mild and 26 with moderate AS) underwent paired CMR and echocardiography at baseline and were followed up with annual echocardiographic examinations for two consecutive years. The baseline characteristics of this cohort are presented in Supplemental Tables 11 and 12. During the follow-up, 14 (28%) patients experienced a progression in AS severity (5 patients with mild AS developed moderate AS, 9 with moderate AS developed severe AS), and 5 required AV replacement. Patients classified into the high-risk group were more likely to experience progression in AS than patients in the low-risk group (Figure 5). Mid-wall LGE was observed in 7 (22%) patients in the high-risk group and 0 (0%) in the low-risk group (p=0.040), and patients with higher LGE-based myocardial fibrosis volumes and indexed extracellular volumes were more likely to be classified into the high-risk group (unadjusted OR: 1.161 [95% CI: 1.070 – 1.280], p=0.002, and 1.451 [95% CI: 1.199 – 1.867], p<0.001, respectively), even after adjusting for the severity of AS (adjusted OR: 1.147 [95% CI: 1.052 – 1.276], p=0.005, and 1.415 [95% CI: 1.161 – 1.825], p=0.002, respectively). When validated in the CMR cohort, the multivariable Cox model developed in the ARIC cohort (Model AS3, Table 4) predicted the progression of AS with a C-index of 0.798 [95% CI: 0.648 – 0.948], and the linear predictor calculated based on this model correlated positively with both the LGE-based myocardial fibrosis volume (r=0.48, p<0.001; Figure 6A) and the indexed extracellular volume (r=0.50, p<0.001; Figure 6B).

**Figure 5.**
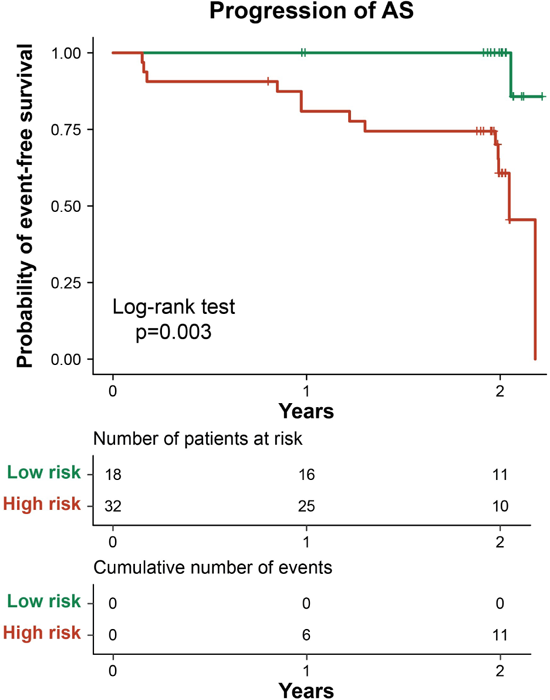
Kaplan-Meier curves showing the event-free survival of patients in the CMR cohort The cut-off value optimized in the ARIC cohort (i.e., the DD probability value of 0.81) was used to stratify patients into low-risk and high-risk groups. CMR – cardiac magnetic resonance; other abbreviations as in Figure 1.

**Figure 6.**
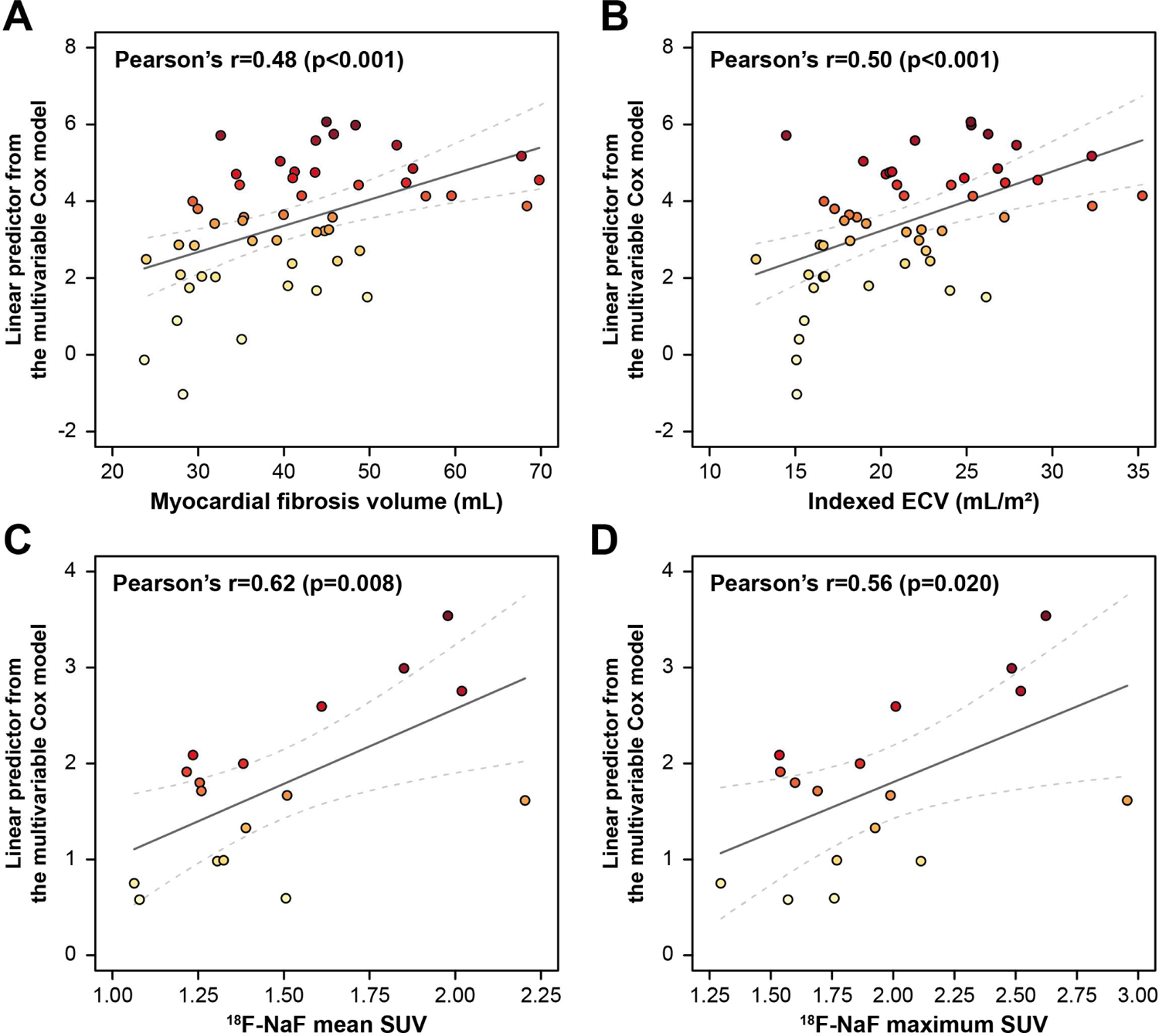
Correlations between the linear predictor calculated based on the multivariable Cox model and LGE-based myocardial fibrosis volume (A), indexed ECV assessed using myocardial T1 mapping (B), and valvular ^18^F-NaF uptake quantified using PET/CT (C and D) ^18^F-NaF – ^18^F-sodium fluoride, ECV – extracellular volume, LGE – late gadolinium enhancement, PET/CT – positron emission tomography combined with computed tomography, SUV – standardized uptake value

#### PET/CT cohort

The clinical characteristics of the PET/CT cohort are shown in Supplemental Table 13. All 18 patients were classified as high-risk. The linear predictor calculated using the final multivariable Cox regression model (Model AS3, Table 4) demonstrated a significant positive correlation with ^18^F-NaF mean SUV (r=0.62, p=0.008; Figure 6C) and maximum SUV (r=0.56, p=0.020; Figure 6D); however, it did not correlate with ^18^F-FDG mean SUV (r=0.15, p=0.558) or maximum SUV (r=0.07, p=0.797). The calculation of the linear predictor is thoroughly described in the Supplemental Results, and examples are provided in Supplemental Table 14.

## DISCUSSION

In 2014, the ACC/AHA valvular heart disease guideline introduced the patient-centered classification of AS, consisting of four stages. The first two stages, referring to early stages of subclinical AV disease, are highly prevalent, as indicated by recent data showing that Stages A (at risk of AS) and B (mild-to-moderate AS) are present in 39% and 17% of older adults, respectively (32). To the best of our knowledge, our current study is the first to investigate the associations between DL-predicted DD and the development and progression of AS in patients with early-stage (i.e., Stage A/B) AV disease (see Central Illustration).

The strength of our study is the utilization of data from various independent cohorts and validation using multimodality imaging. Initially, we applied our previously validated DL model on data from the ARIC cohort study, a population-based prospective study that included individuals from diverse regions of the US. In participants exhibiting AV sclerosis (Stage A), we determined the optimal DD probability threshold (0.81) for identifying individuals with high and low risk of progressing from AV sclerosis to AS. This threshold was subsequently used in an external cohort of patients with mild-to-moderate AS (Stage B) who underwent CMR imaging at baseline and annual echocardiographic assessments for two consecutive years. We found that high-risk patients had larger indexed extracellular volumes on CMR at baseline and were more likely to exhibit a progression in AS during follow-up than those classified into the low-risk group. These findings are consistent with those of previous studies in which a greater extent of diffuse myocardial fibrosis was associated with more severe DD and had additive value for predicting clinical outcomes in patients with AS (36).

Furthermore, in the ARIC cohort, we created a multivariable Cox model incorporating the DL-predicted probability of DD to predict the development of AS from AV sclerosis. The linear predictor calculated based on this multivariable Cox model demonstrated good discriminatory power in predicting the worsening of AS severity in the CMR cohort. Moreover, in another external cohort, the linear predictor was also found to be positively correlated with baseline valvular ^18^F-NaF mean and maximum SUV. Increased valvular calcification activity measured by ^18^F-NaF uptake is known to be present in patients with AV sclerosis and has been shown to be an accurate predictor of disease progression, outperforming all baseline clinical and echocardiographic measures of AS severity (34,37,38). Overall, these findings suggest that the DL-based assessment of DD may identify an underlying milieu of valvular inflammation and calcification and myocardial fibrosis; thus, it can quantify the latent risk associated with the development and progression of AS.

The findings from our study bring to light several unresolved questions regarding myocardial changes in AS. For example, it has been previously believed that the excessive LV afterload in AS induces concentric hypertrophy and DD, and systolic function declines only later when the compensating mechanisms fail to maintain normal wall stress. Nevertheless, multiple studies have recently challenged this long-standing concept, showing that some patients may develop systolic dysfunction before AS becomes severe (39,40). Thus, LV systolic and diastolic dysfunction could occur even at the early stages of calcific AV disease (11). Our findings also support this observation and align with the mounting evidence that suggests mild AV disease (i.e., AV sclerosis and mild AS) occurs in a large proportion of patients with heart failure with preserved EF (HFpEF) (41). Saliency maps and phenome-wide association studies supported links with traditional cardiovascular risk factors and diastolic dysfunction.

Our findings align with observations from a recent study proposing an innovative video-based biomarker for predicting the development and progression of AS (42). This biomarker was found to be correlated with multiple echocardiographic parameters of DD, and activation maps indicated that it also incorporated information from extra-valvular myocardial structures besides the AV (42). Building upon these observations and our study’s findings, it is intriguing to speculate that rather than being sequential phenomena, calcific AV disease and myocardial dysfunction may develop and progress concomitantly, and DD may even precede the development of AS. Previous studies have revealed that calcific AV disease is a highly regulated disease process mediated by several cellular and molecular pathways (43,44). Importantly, many of these pathways also play a central role in the pathophysiology of DD and HFpEF (45,46). For example, inflammation and oxidative stress are major factors contributing to the evolution of AS (47–49). These may be mediated by mitochondrial dysfunction and dysregulation of autophagy that extend even beyond the valve and underlie the development of adverse LV remodeling and DD (12–14). Thus, the identification of DD may capture this underlying systemic milieu of cellular impairment that affects both the valve and the myocardium.

Alternately, beyond being simply coexisting disease entities, it’s also worthwhile speculating whether DD itself creates an environment that accelerates the progression of AV sclerosis. It is well-known that DD is associated with changes in the characteristics of intraventricular blood flow (50–53). Under physiological conditions, the intracavitary flow is redirected and accelerated toward the LV outflow during the pre-ejection period, forming a large anterior vortex and finally interacting with the AV leaflets as they open for LV ejection (50–52). Systolic and diastolic dysfunction disrupt this vortical blood flow (51,52,54), also altering the interaction of the blood flow with the AV. Interestingly, changes in blood flow dynamics have been linked to the accelerated progression of AV sclerosis to AS (55,56). Therefore, another plausible hypothesis could be that such DD-related changes alter shear stress on the AV – a key initiating factor in the development of calcific AV disease (44).

The clinical implications of identifying early-stage AS (Stage A/B) patients who have an increased risk of developing AS and subsequent progression are extraordinary. Although currently, there are no proven pharmacotherapies to prevent or halt the progression of AS, discovering new pathways to mitigate the progression remains a hot topic for research. Multiple clinical trials are underway to investigate the pharmacological prevention of AS (57,58). AV sclerosis represents perhaps the ideal stage for intervention when the disease process is potentially most amenable to intervention; however, the fact that only 10-15% of patients develop AS has precluded trials in this population. The ability of our DL model to identify patients who are predisposed to progression could potentially facilitate candidate selection in clinical trials for pharmacologic interventions addressing the progression and development of AS.

### Limitations

While our study has yielded promising results, it has several limitations that must be acknowledged. First, in our pursuit of complying with current recommendations (27), we opted to revise the upper limit of AV peak velocity from 2.0 to 2.5 m/s for defining AV sclerosis (59). Notably, this adjustment introduces a “gray zone” (AV peak velocity ranging from 2.0 to 2.5 m/s) where inconsistencies exist in guidelines concerning the terminology distinguishing AV sclerosis from mild AS. We recognize that further refinement of risk prediction could be achieved by defining the extent of calcification or valve thickening. Unfortunately, these were not reported during the echocardiographic examination at visit 5 in the ARIC cohort.

Second, it is noteworthy that the ARIC investigators also performed echocardiographic examinations during visit 7. However, the data from these examinations were not made publicly available at the time of our analysis. Therefore, we inferred the progression of AS based on ICD codes, a method with limitations yet frequently resorted to in epidemiological studies (60). Intriguingly, even a previously published study investigating the progression of Stage A/B AV disease based on serial echocardiographic assessment within the ARIC cohort necessitated the use of ICD codes because events such as AV replacements, hospitalizations, death, or being lost to follow-up were frequent (32). To circumvent the unavailability of serial echocardiographic data for the ARIC cohort, we leveraged the validation offered by the independent CMR cohort, where patients underwent three echocardiographic examinations (one at baseline and two at annual follow-up visits), with only 5 patients being lost to follow-up. The consistency of our observations in both the ARIC and CMR cohorts substantiates the validity and generalizability of our DL model.

## CONCLUSIONS

Based on our findings, we conclude that our DL model can efficiently integrate the echocardiographic features of DD and identify the latent risk associated with the development and progression of early-stage AS. Individualized modeling of AV disease trajectories and developing models for predicting disease progression has high clinical relevance for identifying patient subsets who will benefit from risk-mitigation strategies and therapeutic interventions that could potentially reduce the risk of AV disease development and progression.

### CLINICAL PERSPECTIVES

#### COMPETENCY IN MEDICAL KNOWLEDGE

Our DL model can efficiently integrate the echocardiographic features of DD to identify the latent risk associated with the development and progression of early-stage AS. The DL-predicted probability of DD correlated with the extent of myocardial fibrosis quantified using CMR imaging. Moreover, the predictions of the multivariable Cox regression model incorporating the DL-derived probability of DD correlated with valvular ^18^F-NaF uptake assessed by PET/CT, thereby identifying a milieu of valvular inflammation and calcification that is known to be associated with the progression of calcific AV disease.

#### TRANSLATIONAL OUTLOOK

Given its ability to identify patients with early-stage AS who are prone to progression, our DL model has the potential to aid in targeting novel pharmacotherapies to those who are most likely to see a benefit. Thus, by optimizing candidate selection and enabling the timely initiation of pharmacologic treatment, DL-based tools could accelerate the development of long-awaited medical therapies for calcific AV disease.

## ABBREVIATIONS

ARIC: Atherosclerosis Risk in Communities study

AS: aortic stenosis

AV: aortic valve

BioLINCC: Biologic Specimen and Data Repository Information Coordinating Center

CMR: cardiac magnetic resonance

CT: computed tomography

DD: diastolic dysfunction

DL: deep learning

ICD: International Classification of Diseases

LGE: late gadolinium enhancement

PET: positron emission tomography

SUV: standardized uptake value

## Data Availability

All data produced in the present study are available upon reasonable request to the authors.

## Central Illustration

Associations between the DL-predicted DD and the development and progression of AS in patients with early-stage AV disease

In this study, we investigated whether a previously validated echocardiography-based DL model assessing DD could identify the latent risk associated with the progression of early-stage AS. In participants with AV sclerosis from the ARIC cohort study, we determined the optimal DD probability threshold (0.81) for identifying individuals with high and low risk of progressing from AV sclerosis to AS. This threshold was subsequently used in an external cohort of patients with mild-to-moderate AS who underwent CMR imaging at baseline and annual echocardiographic assessments for two consecutive years. We found that high-risk patients (DD probability ≥0.81) had larger indexed extracellular volumes on CMR at baseline and were more likely to exhibit a progression in AS during follow-up than those classified into the low-risk group (DD probability <0.81). Last, in a third cohort with AV sclerosis undergoing PET/CT, the linear predictor calculated using the multivariable Cox model incorporating the DL-predicted probability of DD correlated positively with valvular ^18^F-NaF uptake, confirming that the latent risk identified by the model is associated with the underlying milieu of valvular inflammation and calcification. Abbreviations as in Figures 1, 2, 5, and 6.

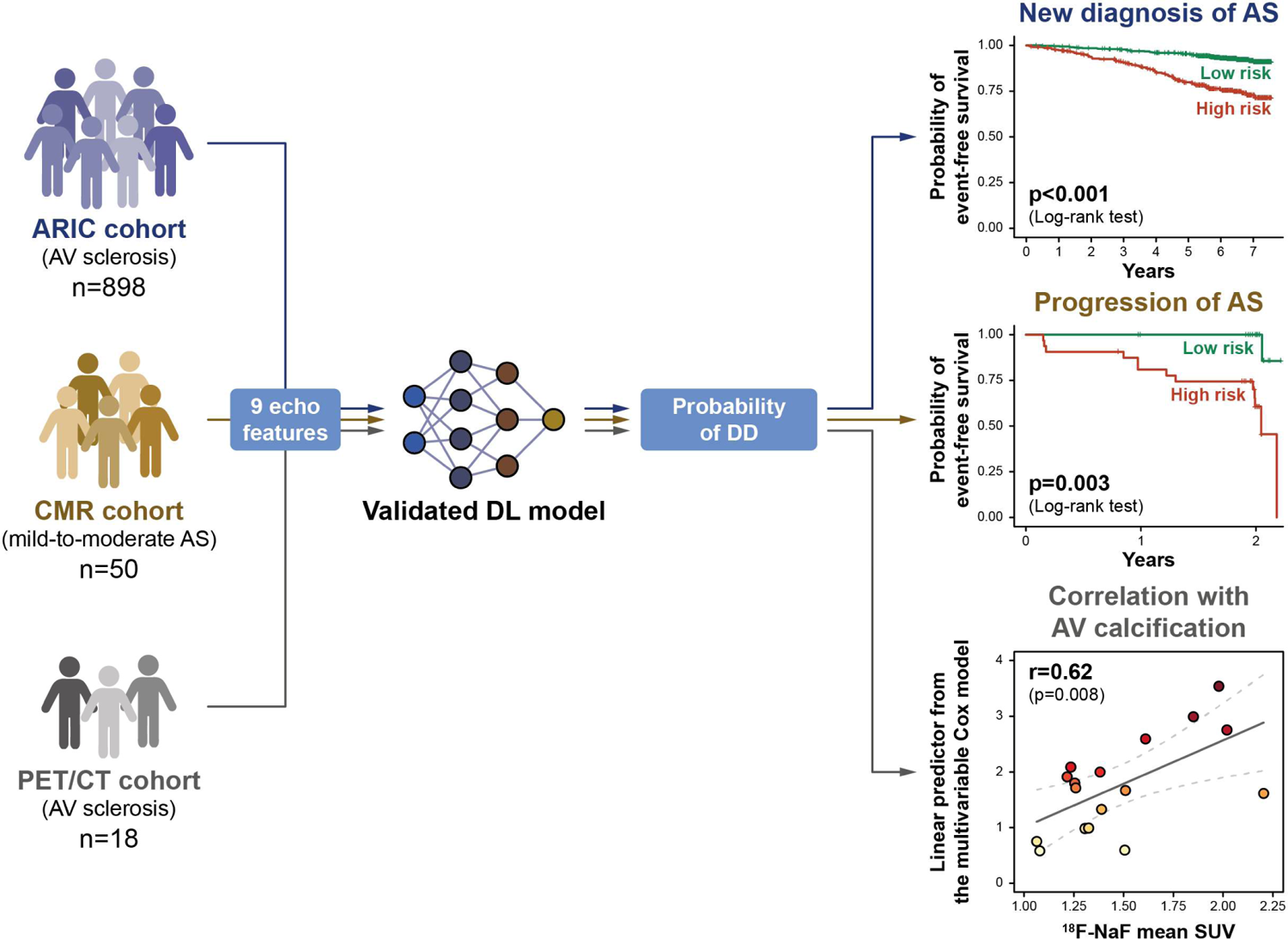

## Notes

FUNDING: The work presented in this paper was supported in part by funds from the National Science Foundation (award number: 1920920). Dr. Tokodi was supported by the New National Excellence Program (ÚNKP-23-4-II-SE-39) of the Ministry of Culture and Innovation in Hungary from the National Research, Development, and Innovation Fund.

CONFLICT OF INTEREST: Dr. Tokodi has received consulting fees from CardioSight outside the submitted work. Dr. Hahn has received speaker fees from Abbott Structural, Baylis Medical, and Edwards Lifesciences; has institutional educational and consulting contracts for which she receives no direct compensation with Abbott Structural, Boston Scientific, Edwards Lifesciences, and Medtronic; and is the chief scientific officer for the Echocardiography Core Laboratory at the Cardiovascular Research Foundation for multiple industry-sponsored trials for which she receives no direct industry compensation. Dr. Dweck is supported by the British Heart Foundation (FS/14/78/31020) and has received the Sir Jules Thorn Award for Biomedical Research 2015 (15/JTA). He has received speaker fees from Pfizer, Radcliffe Cardiology, Bristol Myers Squibb, Edwards, and Novartis and consulting fees from Novartis, Jupiter Bioventures, Beren, and Silence Therapeutics. Dr. Pibarot has received funding from Edwards Lifesciences, Medtronic, Pi-Cardia, and Cardiac Success for echocardiography core laboratory analyses and research studies in transcatheter valve therapies, for which he received no personal compensation. Dr. Pibarot has also received lecture fees from Edwards Lifesciences and Medtronic. Dr. Yanamala serves as an advisor for Turnkey Techstart. Dr. Sengupta serves as an advisor for Echo IQ and RCE Technologies. All other authors have no conflict of interest to declare.

### Competing Interest Statement

Dr. Tokodi has received consulting fees from CardioSight outside the submitted work. Dr. Hahn has received speaker fees from Abbott Structural, Baylis Medical, and Edwards Lifesciences; has institutional educational and consulting contracts for which she receives no direct compensation with Abbott Structural, Boston Scientific, Edwards Lifesciences, and Medtronic; and is the chief scientific officer for the Echocardiography Core Laboratory at the Cardiovascular Research Foundation for multiple industry-sponsored trials for which she receives no direct industry compensation. Dr. Dweck is supported by the British Heart Foundation (FS/14/78/31020) and has received the Sir Jules Thorn Award for Biomedical Research 2015 (15/JTA). He has received speaker fees from Pfizer, Radcliffe Cardiology, Bristol Myers Squibb, Edwards, and Novartis and consulting fees from Novartis, Jupiter Bioventures, Beren, and Silence Therapeutics. Dr. Pibarot has received funding from Edwards Lifesciences, Medtronic, Pi-Cardia, and Cardiac Success for echocardiography core laboratory analyses and research studies in transcatheter valve therapies, for which he received no personal compensation. Dr. Pibarot has also received lecture fees from Edwards Lifesciences and Medtronic. Dr. Yanamala serves as an advisor for Turnkey Techstart. Dr. Sengupta serves as an advisor for Echo IQ and RCE Technologies. All other authors have no conflict of interest to declare.

### Funding Statement

The work presented in this paper was supported in part by funds from the National Science Foundation (award number: 1920920). Dr. Tokodi was supported by the New National Excellence Program (UNKP-23-4-II-SE-39) of the Ministry of Culture and Innovation in Hungary from the National Research, Development, and Innovation Fund.

