## Supplementary Appendix for "Deep Learning Model of Diastolic Dysfunction Risk Stratifies the Progression of Early-Stage Aortic Stenosis"

**SUPPLEMENTAL METHODS**

**Echocardiographic protocol in the ARIC cohort study**

Echocardiographic examinations were performed by experienced echocardiographers at each field center using commercial ultrasound systems (iE33, Philips Medical Systems with Vision 2011). The protocol included 2D, color Doppler, spectral Doppler, and tissue Doppler imaging. All measurements and evaluations were conducted at the Atherosclerosis Risk in Communities (ARIC) Echocardiography Reading Center (Brigham and Women’s Cardiac Imaging Core Laboratory, Boston, MA). All analyses were over-read (blinded to study participants’ clinical information) and were approved by board-certified echocardiographers.

**Covariates of interest in the ARIC cohort**

The covariates of interest were assessed at visit 5 unless specified otherwise. Age, sex, and race (black or white) were assessed at visit 1. Smoking status (current, former, or never) and use of medications (i.e., antidiabetic, antihypertensive, and cholesterol-lowering medications) were obtained from interviewer-administered questionnaires. Diabetes was defined as a fasting glucose level of 126 mg/dL or higher, a non-fasting glucose level of 200 mg/dL or higher, a hemoglobin A1C level of 6.5 % or higher, a self-reported physician diagnosis of diabetes, or the use of antidiabetic medications. Blood pressure was measured three times by a trained technician following a protocol, and the mean of the second and third readings was used in the analysis. Hypertension was defined as a measured blood pressure of 140/90 mmHg or higher or the use of antihypertensive medication. History of coronary heart disease (CHD) and stroke was defined by self-reported history at enrolment (1987 – 1989) or incident cases adjudicated by the ARIC study physician panel. Participants with an electrocardiogram showing atrial fibrillation (AF) or atrial flutter during study visits (as adjudicated by a trained cardiologist) or hospitalization with an AF diagnosis (International Classification of Diseases [ICD]-9 code of 427.3, 427.31, or 427.32 or ICD-10 code of I48 or I48.x at any position) before visit 5 were considered to have a history of AF. Participants with a glomerular filtration rate below 60 mL/min/1.73m^2^ at visit 5 or an event with any ICD code implying chronic kidney disease (CKD) before visit 5 were considered to have CKD (1).

**Development of the DL model for assessing DD**

To develop a deep learning (DL) model for assessing diastolic dysfunction (DD), we first performed topological data analysis (TDA) to simultaneously evaluate nine echocardiographic features (i.e., left ventricular mass index [LVMi], left ventricular ejection fraction [LVEF], early mitral inflow velocity [E], late mitral inflow velocity [A], E/A, early diastolic mitral annular velocity at septal position [e’], E/e’, left atrial volume index [LAVi], and tricuspid regurgitation peak velocity [TRV]) of 1,242 patients with varying degrees of systolic and diastolic dysfunction (2,3). TDA applies the tools of shape analysis to identify and connect highly similar data points (e.g., patients) in a multi-dimensional space and then plots the identified connections as a two-dimensional similarity network (4). The generated network consists of nodes (representing collections of similar data points) connected by edges (i.e., lines between two nodes) if they have at least one data point in common. In our dataset, TDA resulted in a circular similarity network that we divided into two parts based on the differences in the clinical characteristics and outcomes of patients located in its different regions: one including patients with low risk of DD and one encompassing patients with high risk of DD. Thus, we could assign a label to each of the 1,242 patients in the similarity network and train classifiers (i.e., supervised machine learning models), enabling the assessment of DD in new, previously unseen patients.

For model training and evaluation, we used BigML (BigML, Inc., Corvallis, OR, <http://bigml.com>) – a cloud-based machine-learning platform. After splitting the cohort into training and test sets in an 80:20 ratio, we trained different classifiers (i.e., decision trees, ensembles of decision trees, logistic regression models, and deep neural networks) in the training set (with cross-validation) and subsequently evaluated them in the test set. We applied the OptiML optimization process for model selection and hyperparameter tuning, which uses Bayesian hyperparameter optimization to create and evaluate hundreds of supervised models and returns a list of the best-performing ones (5). Then, these models can be used individually or combined into a so-called *fusion model*.

Our DL model is a *fusion model* that combines the seven best-performing deep neural networks and averages their predictions (3). In our model, the most important feature for predicting DD is e’, followed by E/e’, LVMi, LVEF, LAVi, TRV, E, A, and E/A in decreasing order of importance. The model has been externally validated in multiple cohorts, showing that higher DL-predicted DD probability values were associated with elevated LV filling pressures, increased neurohormonal activation, lower exercise capacity, and a higher incidence of adverse cardiac events during follow-up (3).

The DL model is publicly available at <https://wvu-model.herokuapp.com>. A more detailed description of model development and evaluation can be found in our previously published papers (2,3).

**Calculation of the linear predictor**

In the present study, we used the linear predictor calculated from the final multivariable Cox model as a “prognostic index”, as recommended by others (6,7). The linear predictor can be calculated for each patient using the following equation:

$$Linear predictor=\sum_{j} (x_{j}-\bar{x}_{j}) \times\beta_{j}$$

where $x_{j}$ is the value of covariate (i.e., predictor) $j$ for the given patient, $\bar{x}_{j}$ is the mean value of covariate $j$ in the entire cohort from which the Cox regression model was derived, and $\beta_{j}$ is the regression coefficient belonging to covariate $j$.

The linear predictor is relative to the cohort from which the Cox regression model was derived: a linear predictor of >0 indicates a higher risk, whereas a linear predictor of <0 indicates a lower risk compared to a reference person. Each covariate of the reference person is calculated as the mean of the given covariate in the entire cohort.

**SUPPLEMENTAL RESULTS**

**Comparing the prognostic power of the DL-derived predictions with the guideline-based DD grading in the ARIC cohort**

Based on the guidelines of the American Society of Echocardiography (ASE) and the European Association of Cardiovascular Imaging (EACVI), 222 (25%) patients had no DD, 459 (51%) had grade I DD, 84 (9%) had grade II or III DD, and 133 (15%) had indeterminate diastolic function (Supplemental Figure 1). The cumulative number of events for both outcomes increased in a graded fashion from patients with no DD to those with grade II-III DD (Supplemental Figure 1). In contrast, patients with indeterminate diastolic function exhibited similar cumulative event rates as those with grade I DD (Supplemental Figure 1).

Most patients (83%) with grade II or III DD were classified into the high-risk group (Figure 4). In contrast, a substantial reclassification was noted for patients with no and grade I DD, as 14% and 39% were discordantly reclassified into the high-risk group, respectively (Supplemental Figure 1). Moreover, 50% of patients with indeterminate diastolic function were reclassified to the high-risk group (Supplemental Figure 1). Thus, the greatest benefit of our DL model lies in its ability to risk-stratify patients whose diastolic function could not be determined using the guideline-based grading.

The DL-predicted probability of DD exhibited a superior prognostic power over guideline-based grading for both endpoints with a significant improvement in Harrel’s C-index (new diagnosis of AS: p<0.001; composite endpoint: p<0.001), integrated discrimination index (p<0.001 for both endpoints), and net reclassification index (p<0.001 for both endpoints) (Supplemental Figure 2, Supplemental Table 8).

**Calculating the linear predictor from the final multivariable Cox model**

In our final multivariable Cox model (Model AS3, Table 4), the reference patient is a 76-years-old white female with no hypertension, diabetes, history of heart failure (HF), history of CHD, or history of AF, who has an aortic valve (AV) peak velocity of 1.74 m/s and a DL-predicted DD probability of 0.5 (Supplemental Table 14). The linear predictor can be calculated for each patient using the following equation:

$$\boldsymbol{Linear predictor}=\left( \left[ \boldsymbol{age} in years \right]-76.110 \right)\times0.042+\left( \left[ \boldsymbol{sex}: 1 if male, 0 if female \right] - 0 \right)\times0.006+\left( \left[ \boldsymbol{race}: 1 if black, 0 if white \right]-0 \right)\times-1.003+\left( \left[ \boldsymbol{hypertension}:1 if yes, 0 if no \right]-0 \right)\times0.386+\left( \left[ \boldsymbol{diabetes}:1 if yes, 0 if no \right]-0 \right)\times0.388+\left( \left[ \boldsymbol{history of HF}:1 if yes, 0 if no \right]-0 \right)\times0.579+\left( \left[ \boldsymbol{history of CHD}:1 if yes, 0 if no \right]-0 \right)\times0.190+\left( \left[ \boldsymbol{history of AF}: 1 if yes, 0 if no \right]-0 \right)\times0.283+\left( \left[ \boldsymbol{AV peak velocity} in m/s \right]-1.737 \right)\times1.738+(\left[ \boldsymbol{probability of DD} predicted by the DL model \right]-0.500)\times1.248$$

The probability of DD can be calculated at the following website: <https://wvu-model.herokuapp.com>. In Supplemental Table 14, we presented the baseline characteristics of four example patients and their corresponding linear predictors calculated using the equation above.

**Supplemental Table 1** Diagnosis codes used in the current study

|  | **ICD-9** | **ICD-10** |
| --- | --- | --- |
| **Acute myocardial infarction** | 410, 410.x | I21, I21.x, I22, I22.x |
| **Aortic stenosis** | 424.1 | I35.0, I35.2 |
| **Atrial fibrillation** | 427.3, 427.31, 427.32 | I48, I48.x |
| **Congenital heart disease** | 745, 745.x, 746, 746.x | Q20, Q20.x, Q21, Q21.x, Q22, Q22.x, Q24, Q24.x |
| **Heart failure** | 428, 428.x | I50, I50.x |
| **Hypertrophic obstructive cardiomyopathy** | 425.11 | I42.1 |
| **Prosthetic valve** | V42.2, V43.3 | Z95.2, Z95.3, Z95.4 |

To establish the diagnosis of chronic kidney disease, we also used the ICD-9 and ICD-10 diagnosis codes recommended by Grams et al. (1) besides evaluating the glomerular filtration rate at visit 5.

ICD-9 – 9th revision of the International Classification of Diseases, ICD-10 – 10th revision of the International Classification of Diseases

**Supplemental Table 2** Procedure codes used in the current study

|  | **ICD-9** | **ICD-10** |
| --- | --- | --- |
| **Aortic valve interventions** | 35.01, 35.05, 35.06, 35.11, 35.21, 35.22, 35.96 | 027Fx, 02NFx, 02QFx, 02RFx |
| **Echocardiography** | 88.72 | B244YZZ, B244ZZZ,  B245YZZ, B245ZZZ,  B246YZZ, B246ZZZ,  B24BYZZ, B24BZZZ,  B244ZZ4, B245ZZ4,  B246ZZ4, B24BZZ4 |

To establish the diagnosis of chronic kidney disease, we also used the ICD-9 and ICD-10 procedure codes recommended by Grams et al. (1) besides evaluating the glomerular filtration rate at visit 5.

ICD-9 – 9th revision of the International Classification of Diseases, ICD-10 – 10th revision of the International Classification of Diseases

**Supplemental Table 3** R packages used in the current study

| **R package** | **Version** |
| --- | --- |
| **compareC** | 1.3.2 |
| **dplyr** | 1.0.10 |
| **Hmisc** | 4.7-2 |
| **forestplot** | 3.1.1 |
| **nonnestcox** | 0.0.0.9000 |
| **pec** | 2023.4.12 |
| **prodlim** | 2019.11.13 |
| **pspline** | 1.0.19 |
| **smoothHR** | 1.0.4 |
| **survIDINRI** | 1.1.2 |
| **survival** | 3.2.13 |
| **survminer** | 0.4.9 |
| **tidyr** | 1.2.1 |

**Supplemental Table 4** Univariable and multivariable Cox regression models (including laboratory parameters as covariates) for predicting the new diagnosis of AS in the ARIC cohort

|  | **Univariable models** | | **Multivariable model**  (C-index: 0.788, AIC: 1,325) | |
| --- | --- | --- | --- | --- |
|  | **HR [95% CI]** | **P-value** | **HR [95% CI]** | **P-value** |
| Age | 1.082  [1.046 – 1.120] | <0.001 | 1.013  [0.974 – 1.053] | 0.523 |
| Male sex | 1.551  [1.079 – 2.229] | 0.018 | 0.967  [0.619 – 1.510] | 0.883 |
| Black race | 0.397  [0.193 – 0.815] | 0.012 | 0.566  [0.267 – 1.203] | 0.139 |
| Creatinine | 1.434  [1.181 – 1.741] | <0.001 | 0.968  [0.643 – 1.458] | 0.877 |
| Hemoglobin A1C | 1.278  [1.068 – 1.530] | 0.008 | 1.210  [0.995 – 1.473] | 0.057 |
| HDL-C | 0.975  [0.959 – 0.990] | 0.002 | 0.993  [0.975 – 1.010] | 0.413 |
| Log_10_(NT-proBNP) | 5.993  [4.155 – 8.644] | <0.001 | 3.816  [2.275 – 6.402] | <0.001 |
| Log_10_(hs-Troponin T) | 6.048  [3.310 – 11.053] | <0.001 | 0.789  [0.302 – 2.061] | 0.628 |
| RDW | 1.178  [1.004 – 1.382] | 0.045 | 1.116  [0.944 – 1.318] | 0.198 |
| AV peak velocity | 9.915  [5.403 – 18.195] | <0.001 | 4.612  [2.394 – 8.884] | <0.001 |
| DL-predicted probability of DD | 5.399  [3.253 – 8.962] | <0.001 | 2.589  [1.449 – 4.612] | 0.001 |

The multivariable model includes only 846 participants (110 with a new diagnosis of aortic stenosis during follow-up) due to missing values.

AIC – Akaike information criterion, AV – aortic valve, CI – confidence interval, DD – diastolic dysfunction, DL – deep learning, HDL-C – high-density lipoprotein cholesterol, HR – hazard ratio, hs – high-sensitivity, NT-proBNP – N-terminal pro-brain natriuretic peptide, RDW – red cell distribution width
**Supplemental Table 5** Univariable and multivariable Cox regression models (including smoking status, chronic kidney disease, and medications as covariates) for predicting the new diagnosis of AS in the ARIC cohort

|  | **Univariable models** | | **Multivariable model**  (C-index: 0.756, AIC: 1,352) | |
| --- | --- | --- | --- | --- |
|  | **HR [95% CI]** | **P-value** | **HR [95% CI]** | **P-value** |
| Age | 1.082  [1.046 – 1.120] | <0.001 | 1.043  [1.006 – 1.081] | 0.023 |
| Male sex | 1.551  [1.079 – 2.229] | 0.018 | 1.114  [0.749 – 1.658] | 0.594 |
| Black race | 0.397  [0.193 – 0.815] | 0.012 | 0.434  [0.198 – 0.952] | 0.037 |
| Chronic kidney disease | 1.923  [1.338 – 2.764] | <0.001 | 1.478  [1.005 – 2.175] | 0.047 |
| Current smoker | 1.286  [0.577 – 2.867] | 0.538 | 1.654  [0.733 – 3.731] | 0.226 |
| Former smoker | 1.178  [0.794 – 1.748] | 0.416 | 1.026  [0.685 – 1.535] | 0.902 |
| Antihypertensive medications | 2.005  [1.148 – 3.503] | 0.015 | 1.200  [0.651 – 2.210] | 0.559 |
| Antidiabetic medications | 1.688  [1.141 – 2.499] | 0.009 | 1.327  [0.855 – 2.058] | 0.207 |
| Statin | 1.351  [0.927– 1.970] | 0.117 | 1.105  [0.716 – 1.705] | 0.652 |
| AV peak velocity | 9.915  [5.403 – 18.195] | <0.001 | 5.845  [3.077 – 11.100] | <0.001 |
| DL-predicted probability of DD | 5.399  [3.253 – 8.962] | <0.001 | 3.684  [2.152 – 6.307] | <0.001 |

The multivariable model includes only 826 participants (110 with a new diagnosis of aortic stenosis during follow-up) due to missing values.

AIC – Akaike information criterion, AV – aortic valve, CI – confidence interval, DD – diastolic dysfunction, DL – deep learning, HR – hazard ratio, hs – high-sensitivity

**Supplemental Table 6** Sequential multivariable Cox regression models for predicting the composite endpoint in the ARIC cohort

|  | **Model C1**  (C-index: 0.766, AIC: 717) | | **Model C2**  (C-index: 0.817, AIC: 692) | |
| --- | --- | --- | --- | --- |
|  | **HR [95% CI]** | **P-value** | **HR [95% CI]** | **P-value** |
| Age | 1.102  [1.050 – 1.158] | <0.001 | 1.078  [1.026 – 1.132] | 0.003 |
| Male sex | 1.079  [0.637 – 1.830] | 0.777 | 0.973  [0.573 – 1.652] | 0.919 |
| Black race | 3.429  [0.825 – 14.257] | 0.090 | 3.743  [0.895 – 15.658] | 0.071 |
| AV peak velocity | 10.592  [4.425 – 25.351] | <0.001 | 7.998  [3.412 – 18.745] | <0.001 |
| DL-predicted probability of DD |  |  | 7.033  [3.036 – 16.290] | <0.001 |

AIC – Akaike information criterion, AV – aortic valve, CI – confidence interval, DL – deep learning, HR – hazard ratio

**Supplemental Table 7** Results of the sensitivity analysis in the ARIC cohort

|  | **Univariable model** | | **Multivariable model** | |
| --- | --- | --- | --- | --- |
|  | **HR [95% CI]** | **P-value** | **HR [95% CI]** | **P-value** |
| New diagnosis of AS – based on echocardiography and/or ICD codes | 5.399  [3.253 – 8.962] | <0.001 | 3.482  [2.061 – 5.884] | <0.001 |
| New diagnosis of AS – ascertained by echocardiography | 8.522  [4.161 – 17.460] | <0.001 | 4.815  [2.323 – 9.980] | <0.001 |

AS – aortic stenosis, CI – confidence interval, ICD – International Classification of Diseases, HR – hazard ratio

**Supplemental Table 8** Comparing the prognostic value of the guideline-based DD grading and the DL-predicted probability in the ARIC cohort

|  | **PLR** | **Change in C-index** | | **IDI** | | **NRI (continuous)** | |
| --- | --- | --- | --- | --- | --- | --- | --- |
|  | P-value | Δ | P-value | Estimate | P-value | Estimate | P-value |
| **New diagnosis of AS** | 0.061 | +0.117 | <0.001 | 0.255  [0.147 – 0.359] | <0.001 | 0.504  [0.424 – 0.725] | <0.001 |
| **Composite endpoint** | 0.013 | +0.160 | <0.001 | 0.149  [0.028 – 0.244] | <0.001 | 0.741  [0.426 – 0.831] | <0.001 |

AV – aortic valve, DL – deep learning, IDI – integrated discrimination improvement, NRI – net reclassification improvement, PLR – partial likelihood ratio test

**Supplemental Table 9** Comparing the sequential multivariable Cox regression models for predicting the new diagnosis of AS in the ARIC cohort

|  | **LR** | **Change in C-index** | | **IDI** | | **NRI (continuous)** | |
| --- | --- | --- | --- | --- | --- | --- | --- |
|  | P-value | Δ | P-value | Estimate | P-value | Estimate | P-value |
| **Model AS2 vs. AS1** | <0.001 | +0.038 | 0.009 | 0.037  [0.002 – 0.080] | 0.027 | -0.457  [-0.545 – 0.474] | 0.073 |
| **Model AS3 vs. AS2** | <0.001 | +0.026 | 0.039 | 0.057  [0.022 – 0.095] | <0.001 | 0.688  [0.585 – 0.747] | <0.001 |

Model AS1: clinical variables.

Model AS2: clinical variables + AV peak velocity.

Model AS3: clinical variables + AV peak velocity + DL-predicted probability.

Clinical variables include age, sex, race, hypertension, diabetes, history of heart failure, history of coronary heart disease, and history of atrial fibrillation.

AV – aortic valve, DL – deep learning, IDI – integrated discrimination improvement, LR – likelihood ratio test, NRI – net reclassification improvement

**Supplemental Table 10** Comparing the sequential multivariable Cox regression models for predicting the composite endpoint in the ARIC cohort

|  | **LR** | **Change in C-index** | | **IDI** | | **NRI (continuous)** | |
| --- | --- | --- | --- | --- | --- | --- | --- |
|  | P-value | Δ | P-value | Estimate | P-value | Estimate | P-value |
| **Model C2 vs. C1** | <0.001 | +0.051 | 0.015 | 0.070  [0.027 – 0.116] | <0.001 | 0.778  [0.657 – 0.870] | 0.007 |

Model C1: age + sex + race + AV peak velocity.

Model C2: age + sex + race + AV peak velocity + DL-predicted probability.

AV – aortic valve, DL – deep learning, IDI – integrated discrimination improvement, LR – likelihood ratio test, NRI – net reclassification improvement

**Supplemental Table 11** Baseline characteristics of patients with progression and no progression of AS within the CMR cohort

|  | **Missing**  n (%) | **All patients**  n=50 | **Progression of AS**  n=14 | **No proregression of AS**  n=36 | **P-value** |
| --- | --- | --- | --- | --- | --- |
| **Demographics, vitals, risk factors** | | | | | |
| Age, years | 0 (0) | 68 (59 – 75) | 69 (64 – 75) | 68 (56 – 73) | 0.256 |
| Male sex | 0 (0) | 36 (72) | 9 (64) | 27 (75) | 0.684 |
| Black race | 0 (0) | 0 (0) | 0 (0) | 0 (0) | - |
| BMI, kg/m^2^ | 0 (0) | 27.8 (26.2 – 31.2) | 26.5 (25.0 – 31.1) | 28.0 (27.2 – 31.4) | 0.217 |
| BSA, m^2^ | 0 (0) | 1.92 ± 0.19 | 1.90 ± 0.18 | 1.93 ± 0.19 | 0.599 |
| SBP, mmHg | 0 (0) | 149 ± 22 | 147 ± 20 | 149 ± 23 | 0.733 |
| DBP, mmHg | 0 (0) | 84 ± 12 | 84 ± 11 | 84 ± 12 | 0.985 |
| HR, 1/min | 0 (0) | 63 ± 11 | 65 ± 11 | 62 ± 11 | 0.375 |
| Hypertension | 0 (0) | 33 (66) | 11 (79) | 22 (61) | 0.327 |
| Diabetes | 0 (0) | 9 (18) | 0 (0) | 9 (25) | 0.047 |
| History of HF | 0 (0) | 0 (0) | 0 (0) | 0 (0) | - |
| History of CHD | 0 (0) | 17 (34) | 6 (43) | 11 (31) | 0.623 |
| History of AF | 0 (0) | 0 (0) | 0 (0) | 0 (0) | - |
| **Medications** | | | | | |
| ACE-I or ARB | 0 (0) | 22 (44) | 8 (57) | 14 (39) | 0.395 |
| Beta-blockers | 0 (0) | 15 (30) | 1 (7) | 14 (39) | 0.039 |
| Diuretics | 0 (0) | 18 (36) | 8 (57) | 10 (28) | 0.106 |
| Statin | 0 (0) | 21 (42) | 5 (36) | 16 (44) | 0.752 |
| **Laboratory results** | | | | | |
| Creatinine, mg/dL | 1 (2) | 0.86 (0.80 – 0.95) | 0.81 (0.77 – 0.91) | 0.87 (0.81 – 0.95) | 0.141 |
| GFR, mL/min/1.73m^2^ | 1 (2) | 81 (72 – 93) | 75 (72 – 91) | 86 (69 – 93) | 0.894 |
| BNP, pg/mL | 3 (6) | 16.6 (5.2 – 42.1) | 26.5 (11.5 – 53.8) | 16.1 (5.0 – 27.6) | 0.170 |
| hs-Troponin I, ng/mL | 1 (2) | 4.5 (3.1 – 8.9) | 8.2 (4.0 – 14.0) | 4.1 (3.0 – 6.6) | 0.056 |
| **Echocardiographic parameters** | | | | | |
| IVSd, cm | 1 (2) | 1.30 (1.20 – 1.40) | 1.40 (1.30 – 1.40) | 1.3 (1.20 – 1.40) | 0.125 |
| LVPWd, cm | 1 (2) | 1.30 (1.20 – 1.40) | 1.35 (1.30 – 1.40) | 1.3 (1.10 – 1.40) | 0.167 |
| LVIDd, cm | 1 (2) | 4.41 ± 0.51 | 4.34 ± 0.59 | 4.44 ± 0.48 | 0.537 |
| LVIDs, cm | 2 (4) | 3.12 ± 0.43 | 3.09 ± 0.56 | 3.13 ± 0.38 | 0.735 |
| LVMi, g/m^2^ | 1 (2) | 113.0 ± 29.8 | 118.1 ± 29.3 | 111.0 ± 30.1 | 0.454 |
| LVEF, % | 2 (4) | 56.5 ± 7.4 | 56.0 ± 8.3 | 56.7 ± 7.2 | 0.776 |
| LA diameter (AP), cm | 2 (4) | 3.94 ± 0.70 | 4.02 ± 0.72 | 3.91 ± 0.70 | 0.643 |
| AV peak velocity m/s | 0 (0) | 3.02 ± 0.56 | 3.37 ± 0.50 | 2.88 ± 0.52 | 0.004 |
| AV mean gr., mmHg | 0 (0) | 20.0 (13.8 – 23.9) | 25.0 (19.5 – 34.2) | 16.8 (12.2 – 22.3) | 0.006 |
| AV area, cm^2^ | 0 (0) | 1.20 (1.08 – 1.46) | 1.12 (1.05 – 1.21) | 1.28 (1.10 – 1.59) | 0.019 |
| Dimensionless index | 0 (0) | 0.36 ± 0.08 | 0.33 ± 0.06 | 0.38 ± 0.09 | 0.094 |
| E, cm/s | 0 (0) | 73 (64 – 82) | 76 (68 – 85) | 71 (59 – 79) | 0.145 |
| E/A | 0 (0) | 0.9 ± 0.3 | 0.8 ± 0.2 | 1.0 ± 0.4 | 0.047 |
| e’ (septal), cm/s | 0 (0) | 5.7 ± 1.8 | 5.2 ± 0.9 | 5.9 ± 2.0 | 0.126 |
| E/e’ (septal) | 0 (0) | 12.9 (10.0 – 16.1) | 16.1 (13.0 – 16.8) | 12.2 (9.1 – 14.4) | 0.021 |
| **CMR imaging parameters** | | | | | |
| LVEDVi, mL/m^2^ | 0 (0) | 72.2 ± 13.5 | 71.9 ± 14.1 | 72.3 ± 13.5 | 0.935 |
| LVESVi, mL/m^2^ | 0 (0) | 23.2 (18.0 – 26.8) | 22.3 (18.0 – 24.0) | 23.5 (17.8 – 27.0) | 0.681 |
| LVEF, % | 0 (0) | 67.8 (63.8 – 70.3) | 68.2 (64.3 – 69.8) | 67.6 (62.9 – 70.6) | 0.983 |
| Longitud. function, mm | 0 (0) | 13.8 ± 2.4 | 14.1 ± 2.0 | 13.7 ± 2.6 | 0.538 |
| LVMi, g/m^2^ | 0 (0) | 82.4 ± 17.1 | 85.5 ± 19.0 | 81.2 ± 16.4 | 0.427 |
| LAVi, mL/m^2^ | 7 (14) | 28.4 (23.1 – 37.7) | 29.3 (24.4 – 37.0) | 28.1 (23.0 – 40.7) | 0.880 |
| Mid-wall LGE | 0 (0) | 7 (14) | 4 (29) | 3 (8) | 0.085 |
| Fibrosis vol. (LGE), mL | 0 (0) | 41.4 ± 11.2 | 43.3 ± 11.1 | 40.6 ± 11.3 | 0.457 |
| Native myocard. T1, ms | 1 (2) | 1,175 ± 35 | 1,187 ± 37.76 | 1,170 ± 33 | 0.125 |
| Partition coefficient | 0 (0) | 0.46 ± 0.03 | 0.48 ± 0.03 | 0.46 ± 0.03 | 0.016 |
| ECV fraction, % | 0 (0) | 27.3 ± 2.4 | 28.0 ± 1.9 | 27.0 ± 2.5 | 0.192 |
| Indexed ECV, mL/m^2^ | 0 (0) | 21.4 ± 5.1 | 22.8 ± 5.1 | 20.9 ± 5.1 | 0.262 |

**Supplemental Table 11** Continued

|  | **Missing**  n (%) | **All patients**  n=50 | **Progression of AS**  n=14 | **No proregression of AS**  n=36 | **P-value** |
| --- | --- | --- | --- | --- | --- |
| **DL-predicted probability of DD** | 0 (0) | 0.94 (0.63 – 0.98) | 0.95 (0.93 – 0.98) | 0.82 (0.39 – 0.98) | 0.064 |
| **DL-predicted high risk** | 0 (0) | 32 (64) | 13 (93) | 19 (53) | 0.009 |

Categorical variables are presented as n (%), and continuous variables as median (interquartile range) or mean ± standard deviation, as appropriate. Comparisons between patients with and without outcomes were performed using unpaired Student’s t-test or Mann-Whitney U test for continuous variables, Chi-squared or Fisher’s exact test for categorical variables, as appropriate.

A – late mitral inflow velocity, ACE-I – angiotensin-converting enzyme inhibitor, AF – atrial fibrillation, AP – anteroposterior, ARB – angiotensin II receptor blocker, AS – aortic stenosis, AV – aortic valve, BMI – body mass index, BNP – brain natriuretic peptide, BSA – body surface area, CHD – coronary heart disease, CMR – cardiac magnetic resonance, DBP – diastolic blood pressure, DD – diastolic dysfunction, DL – deep learning, E – early mitral inflow velocity, ECV – extracellular volume, e’ – early diastolic mitral annular velocity, GFR – glomerular filtration rate, HF – heart failure, HR – heart rate, hs – high-sensitivity, IVSd – thickness of the interventricular septum at end-diastole, LA – left atrial, LAVi – left atrial volume index, LGE – late gadolinium enhancement, LVEDVi – left ventricular end-diastolic volume index, LVEF – left ventricular ejection fraction, LVESVi – left ventricular end-systolic volume index, LVIDd – left ventricular internal diameter at end-diastole, LVIDs – left ventricular internal diameter at end-systole, LVMi – left ventricular mass index, LVPWd – thickness of the left ventricular posterior wall at end-diastole, SBP – systolic blood pressure

**Supplemental Table 12** Baseline characteristics of the high-risk and low-risk groups within the CMR cohort

|  | **Missing**  n (%) | **All patients**  n=50 | **High-risk patients**  n=32 | **Low-risk patients**  n=18 | **P-value** |
| --- | --- | --- | --- | --- | --- |
| **Demographics, vitals, risk factors** | | | | | |
| Age, years | 0 (0) | 68 (59 – 75) | 71 (65 – 75) | 60 (51 – 67) | <0.001 |
| Male sex | 0 (0) | 36 (72) | 25 (78) | 11 (61) | 0.338 |
| Black race | 0 (0) | 0 (0) | 0 (0) | 0 (0) | - |
| BMI, kg/m^2^ | 0 (0) | 27.8 (26.2 – 31.2) | 28.0 (26.0 – 31.2) | 27.7 (27.2 – 31.8) | 0.726 |
| BSA, m^2^ | 0 (0) | 1.92 ± 0.19 | 1.93 ± 0.19 | 1.91 ± 0.18 | 0.635 |
| SBP, mmHg | 0 (0) | 149 ± 22 | 152 ± 20 | 143 ± 24 | 0.149 |
| DBP, mmHg | 0 (0) | 84 ± 12 | 82 ± 10 | 87 ± 14 | 0.166 |
| HR, 1/min | 0 (0) | 63 ± 11 | 61 ± 11 | 65 ± 10 | 0.222 |
| Hypertension | 0 (0) | 33 (66) | 25 (78) | 8 (44) | 0.036 |
| Diabetes | 0 (0) | 9 (18) | 6 (19) | 3 (17) | 1.000 |
| History of HF | 0 (0) | 0 (0) | 0 (0) | 0 (0) | - |
| History of CHD | 0 (0) | 17 (34) | 16 (50) | 1 (6) | 0.002 |
| History of AF | 0 (0) | 0 (0) | 0 (0) | 0 (0) | - |
| **Medications** | | | | | |
| ACE-I or ARB | 0 (0) | 22 (44) | 17 (53) | 5 (28) | 0.151 |
| Beta-blockers | 0 (0) | 15 (30) | 13 (41) | 2 (11) | 0.052 |
| Diuretics | 0 (0) | 18 (36) | 14 (44) | 4 (22) | 0.219 |
| Statin | 0 (0) | 21 (42) | 16 (50) | 5 (28) | 0.219 |
| **Laboratory results** | | | | | |
| Creatinine, mg/dL | 1 (2) | 0.86 (0.80 – 0.95) | 0.88 (0.81 – 0.99) | 0.81 (0.79 – 0.87) | 0.108 |
| GFR, mL/min/1.73m^2^ | 1 (2) | 82 ± 15 | 79 ± 16 | 86 ± 14 | 0.162 |
| BNP, pg/mL | 3 (6) | 16.6 (5.2 – 42.1) | 26.9 (13.2 – 53.9) | 5.2 (5.0 – 16.2) | <0.001 |
| hs-Troponin I, ng/mL | 1 (2) | 4.5 (3.1 – 8.9) | 6.4 (4.1 – 12.4) | 3 (1.8 – 4.0) | <0.001 |
| **Echocardiographic parameters** | | | | | |
| IVSd, cm | 1 (2) | 1.30 (1.20 – 1.40) | 1.4 (1.30 – 1.40) | 1.20 (1.00 – 1.30) | <0.001 |
| LVPWd, cm | 1 (2) | 1.30 (1.20 – 1.40) | 1.4 (1.30 – 1.42) | 1.10 (1.00 – 1.30) | <0.001 |
| LVIDd, cm | 1 (2) | 4.41 ± 0.51 | 4.49 ± 0.47 | 4.28 ± 0.56 | 0.173 |
| LVIDs, cm | 2 (4) | 3.12 ± 0.43 | 3.17 ± 0.43 | 3.02 ± 0.43 | 0.246 |
| LVMi, g/m^2^ | 1 (2) | 109.2  (87.9 – 132.9) | 118.0  (107.0 – 139.7) | 83.0  (81.3 – 103.2) | <0.001 |
| LVEF, % | 2 (4) | 56.5 ± 7.4 | 56.0 ± 8.3 | 57.4 ± 5.5 | 0.569 |
| LA diameter (AP), cm | 2 (4) | 3.94 ± 0.70 | 4.12 ± 0.72 | 3.58 ± 0.52 | 0.011 |
| AV peak velocity m/s | 0 (0) | 3.02 ± 0.56 | 3.18 ± 0.52 | 2.73 ± 0.52 | 0.005 |
| AV mean gr., mmHg | 0 (0) | 20.0 (13.8 – 23.9) | 22.0 (16.8 – 26.4) | 14.3 (11.6 – 19.1) | 0.008 |
| AV area, cm^2^ | 0 (0) | 1.20 (1.08 – 1.46) | 1.17 (1.08 – 1.32) | 1.31 (1.10 – 1.68) | 0.135 |
| Dimensionless index | 0 (0) | 0.35 (0.29 – 0.42) | 0.32 (0.29 – 0.39) | 0.43 (0.34 – 0.46) | 0.014 |
| E, cm/s | 0 (0) | 73 ± 18 | 74 ± 19 | 71 ± 17 | 0.605 |
| E/A | 0 (0) | 0.8 (0.7 – 1.1) | 0.8 (0.7 – 1.0) | 0.9 (0.7 – 1.3) | 0.265 |
| e’ (septal), cm/s | 0 (0) | 5.7 ± 1.8 | 4.9 ± 1.2 | 7.1 ± 1.9 | <0.001 |
| E/e’ (septal) | 0 (0) | 12.9 (10.0 – 16.1) | 14.6 (12.6 – 18.6) | 9.6 (8.4 – 12.0) | <0.001 |
| **CMR imaging parameters** | | | | | |
| LVEDVi, mL/m^2^ | 0 (0) | 69.5 (61.8 – 79.5) | 72.5 (67.0 – 84.4) | 63.5 (57.0 – 73.5) | 0.025 |
| LVESVi, mL/m^2^ | 0 (0) | 23.2 (18.0 – 26.8) | 23.74 (18.0 – 27.5) | 22.5 (17.3 – 25.0) | 0.584 |
| LVEF, % | 0 (0) | 68.0 ± 7.2 | 68.4 ± 8.2 | 67.3 ± 4.9 | 0.557 |
| Longitud. function, mm | 0 (0) | 13.8 ± 2.4 | 13.4 ± 2.4 | 14.4 ± 2.3 | 0.162 |
| LVMi, g/m^2^ | 0 (0) | 82.5 (67.5 – 93.0) | 88.5 (78.0 – 96.8) | 68.0 (60.3 – 81.0) | <0.001 |
| LAVi, mL/m^2^ | 7 (14) | 32.1 ± 13.0 | 35.2 ± 13.9 | 26.4 ± 8.7 | 0.032 |
| Mid-wall LGE | 0 (0) | 7 (14) | 7 (22) | 0 (0) | 0.040 |
| Fibrosis vol. (LGE), mL | 0 (0) | 41.4 ± 11.2 | 45.6 ± 10.8 | 33.9 ± 7.8 | <0.001 |
| Native myocard. T1, ms | 1 (2) | 1,175 ± 35 | 1,185 ± 33 | 1,156 ± 31 | 0.005 |
| Partition coefficient | 0 (0) | 0.46 ± 0.03 | 0.47 ± 0.04 | 0.45 ± 0.02 | 0.006 |
| ECV fraction, % | 0 (0) | 27.3 ± 2.4 | 28.0 ± 2.5 | 26.0 ± 1.7 | 0.004 |
| Indexed ECV, mL/m^2^ | 0 (0) | 21.1 (16.9 – 25.2) | 22.5 (20.6 – 26.4) | 16.6 (15.6 – 18.8) | <0.001 |

**Supplemental Table 12** Continued

|  | **Missing**  n (%) | **All patients**  n=50 | **High-risk patients**  n=32 | **Low-risk patients**  n=18 | **P-value** |
| --- | --- | --- | --- | --- | --- |
| **DL-predicted probability of DD** | 0 (0) | 0.94 (0.63 – 0.98) | 0.97 (0.95 – 0.98) | 0.35 (0.11 – 0.67) | <0.001 |
| **DL-predicted high risk** | 0 (0) | 32 (64) | 32 (100) | 0 (0) | <0.001 |

Categorical variables are presented as n (%), and continuous variables as median (interquartile range) or mean ± standard deviation, as appropriate. Comparisons between patients with and without outcomes were performed using unpaired Student’s t-test or Mann-Whitney U test for continuous variables, Chi-squared or Fisher’s exact test for categorical variables, as appropriate.

A – late mitral inflow velocity, ACE-I – angiotensin-converting enzyme inhibitor, AF – atrial fibrillation, AP – anteroposterior, ARB – angiotensin II receptor blocker, AS – aortic stenosis, AV – aortic valve, BMI – body mass index, BNP – brain natriuretic peptide, BSA – body surface area, CHD – coronary heart disease, CMR – cardiac magnetic resonance, DBP – diastolic blood pressure, DD – diastolic dysfunction, DL – deep learning, E – early mitral inflow velocity, ECV – extracellular volume, e’ – early diastolic mitral annular velocity, GFR – glomerular filtration rate, HF – heart failure, HR – heart rate, hs – high-sensitivity, IVSd – thickness of the interventricular septum at end-diastole, LA – left atrial, LAVi – left atrial volume index, LGE – late gadolinium enhancement, LVEDVi – left ventricular end-diastolic volume index, LVEF – left ventricular ejection fraction, LVESVi – left ventricular end-systolic volume index, LVIDd – left ventricular internal diameter at end-diastole, LVIDs – left ventricular internal diameter at end-systole, LVMi – left ventricular mass index, LVPWd – thickness of the left ventricular posterior wall at end-diastole, SBP – systolic blood pressure

**Supplemental Table 13** Baseline clinical and imaging characteristics of the PET/CT cohort

|  | **Missing**  n (%) | **PET/CT cohort**  n=18 |
| --- | --- | --- |
| **Demographics, vitals, risk factors** |  |  |
| Age, years | 0 (0) | 74 (68 – 78) |
| Male sex | 0 (0) | 14 (78) |
| BMI, kg/m^2^ | 0 (0) | 27.3 (24.2 – 30.3) |
| BSA, m^2^ | 0 (0) | 1.94 (1.85 – 2.08) |
| SBP, mmHg | 0 (0) | 148 (132 – 165) |
| DBP, mmHg | 0 (0) | 83 (77 – 87) |
| HR, 1/min | 0 (0) | 68 (60 – 75) |
| Hypertension | 0 (0) | 10 (56) |
| Diabetes | 0 (0) | 3 (17) |
| Smoking status | 0 (0) |  |
| Never smoker |  | 11 (61) |
| Current smoker |  | 2 (11) |
| Former smoker |  | 5 (28) |
| History of heart failure | 0 (0) | 3 (17) |
| History of coronary heart disease | 0 (0) | 8 (44) |
| History of atrial fibrillation | 0 (0) | 7 (39) |
| History of stroke | 0 (0) | 1 (6) |
| **Medications** |  |  |
| Antihypertensive medications | 0 (0) | 15 (83) |
| Antidiabetic medications | 0 (0) | 1 (6) |
| Statin | 0 (0) | 10 (56) |
| Lipid-lowering medications | 0 (0) | 1 (6) |
| **Echocardiographic parameters** |  |  |
| IVSd, cm | 0 (0) | 1.34 (1.20 – 1.40) |
| LVPWd, cm | 0 (0) | 1.30 (1.12 – 1.40) |
| LVIDd, cm | 0 (0) | 4.60 (4.10 – 5.20) |
| LVIDs, cm | 0 (0) | 3.35 (3.10 – 4.07) |
| LVMi, g/m^2^ | 0 (0) | 117.0 (100.2 – 151.7) |
| LVEF, % | 0 (0) | 60.0 (54.0 – 60.0) |
| LAVi, mL/m^2^ | 3 (17) | 32.8 (22.6 – 45.6) |
| AV peak velocity, m/s | 0 (0) | 1.70 (1.58 – 1.80) |
| E, cm/s | 0 (0) | 67 (61 – 86) |
| E/A | 3 (17) | 0.79 (0.70 – 1.01) |
| e’ (septal), cm/s | 2 (11) | 5.6 (4.6 – 6.8) |
| E/e’ (septal) | 2 (11) | 12.6 (10.4 – 16.8) |
| TRV, m/s | 12 (67) | 2.40 (2.34 – 2.60) |
| **Non-contrast CT parameters** |  |  |
| Agatston AV calcium score, AU | 0 (0) | 271 (70 – 562) |
| **^18^F-NaF PET parameters** |  |  |
| Mean SUV | 1 (6) | 1.38 (1.25 – 1.61) |
| Maximum SUV | 1 (6) | 1.86 (1.60 – 2.11) |
| **^18^F-FDG PET parameters** |  |  |
| Mean SUV | 1 (6) | 1.74 (1.46 – 1.88) |
| Maximum SUV | 1 (6) | 1.93 (1.71 – 2.07) |

Continuous variables are presented as median (interquartile range) and categorical variables as n (%).

^18^F-NaF – ^18^F-sodium fluoride, ^18^F-FDG – ^18^F-fluorodeoxyglucose, A – late mitral inflow velocity, AS – aortic stenosis, AV – aortic valve, BMI – body mass index, BSA – body surface area, DBP – diastolic blood pressure, E – early mitral inflow velocity, e’ – early diastolic mitral annular velocity, HR – heart rate, IVSd – thickness of the interventricular septum at end-diastole, LAVi – left atrial volume index, LVEF – left ventricular ejection fraction, LVIDd – left ventricular internal diameter at end-diastole, LVIDs – left ventricular internal diameter at end-systole, LVMi – left ventricular mass index, LVPWd – thickness of the left ventricular posterior wall at end-diastole, PET – positron emission tomography, SBP – systolic blood pressure, SUV – standardized uptake value, TRV –tricuspid regurgitation peak velocity

**Supplemental Table 14** Baseline characteristics and linear predictors of four example patients and the reference patient

|  | **Example patient #1** | **Example patient #2** | **Reference patient** | **Example patient #3** | **Example patient #4** |
| --- | --- | --- | --- | --- | --- |
| Age, years | 71 | 31 | 76.110 | 82 | 79 |
| Sex | male | female | female | male | male |
| Race | black | white | white | white | white |
| Hypertension | no | no | no | yes | yes |
| Diabetes | no | no | no | yes | no |
| History of HF | no | no | no | yes | no |
| History of CHD | no | no | no | yes | yes |
| History of AF | no | no | no | no | no |
| AV peak velocity, m/s | 1.5 | 1.9 | 1.737 | 2.5 | 3.9 |
| DL-predicted probability of DD | 0.008 | 0.162 | 0.500 | 1.000 | 0.983 |
| **Linear predictor** | **-2.238** | **-2.033** | **0.000** | **3.746** | **5.065** |

The linear predictor was calculated using the equation in the Supplemental Results (see the *Calculating the linear predictor from the final multivariable Cox model* subsection).

AF – atrial fibrillation, AV – aortic valve, CHD – coronary heart disease, DD – diastolic dysfunction, DL – deep learning, HF – heart failure

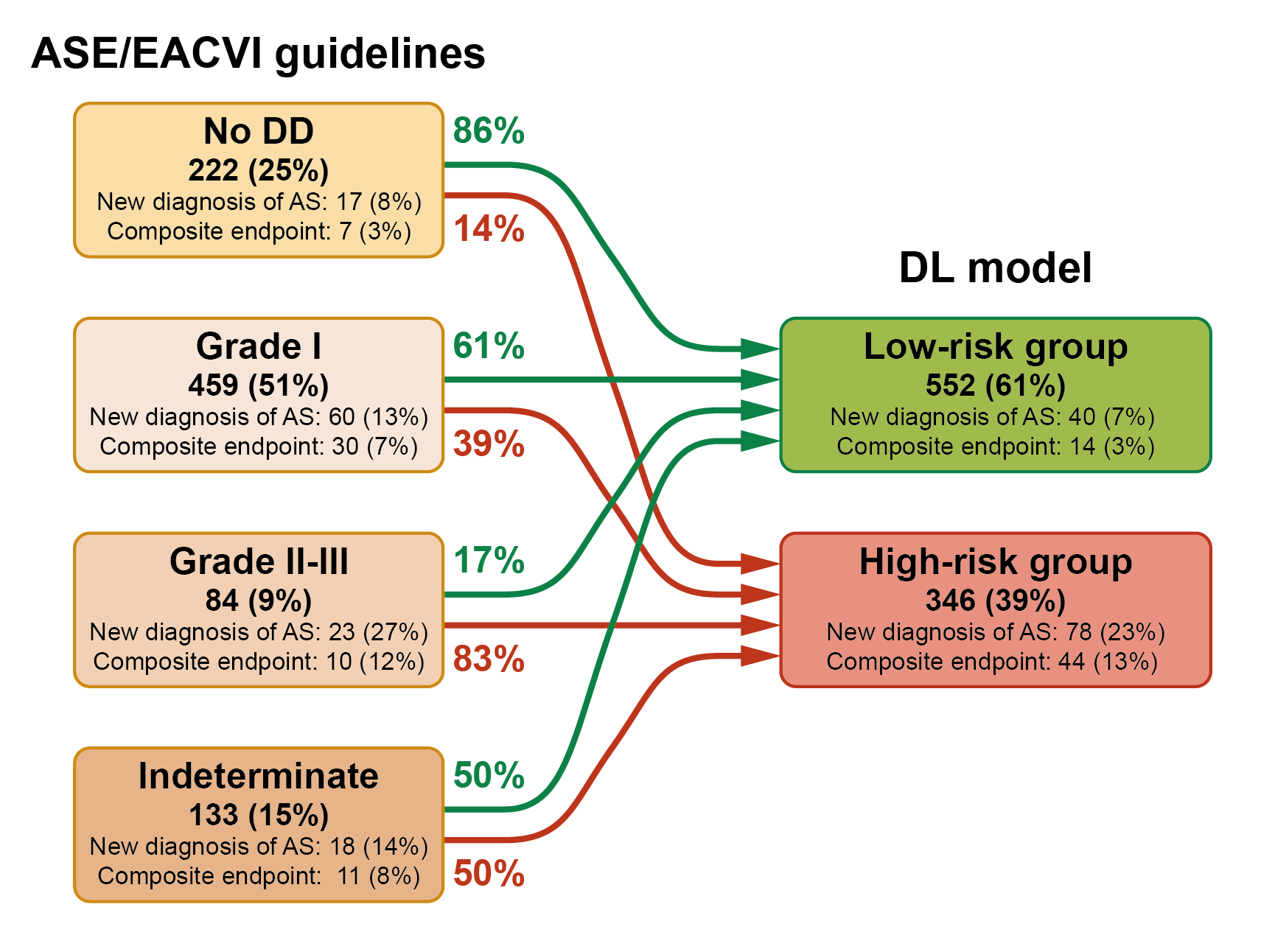

**Supplemental Figure 1** Reclassification of the guideline-based DD grades to DL-derived risk groups in the ARIC cohort

AS – aortic stenosis, ASE – American Society of Echocardiography, DD – diastolic dysfunction, DL – deep learning, EACVI – European Association of Cardiovascular Imaging

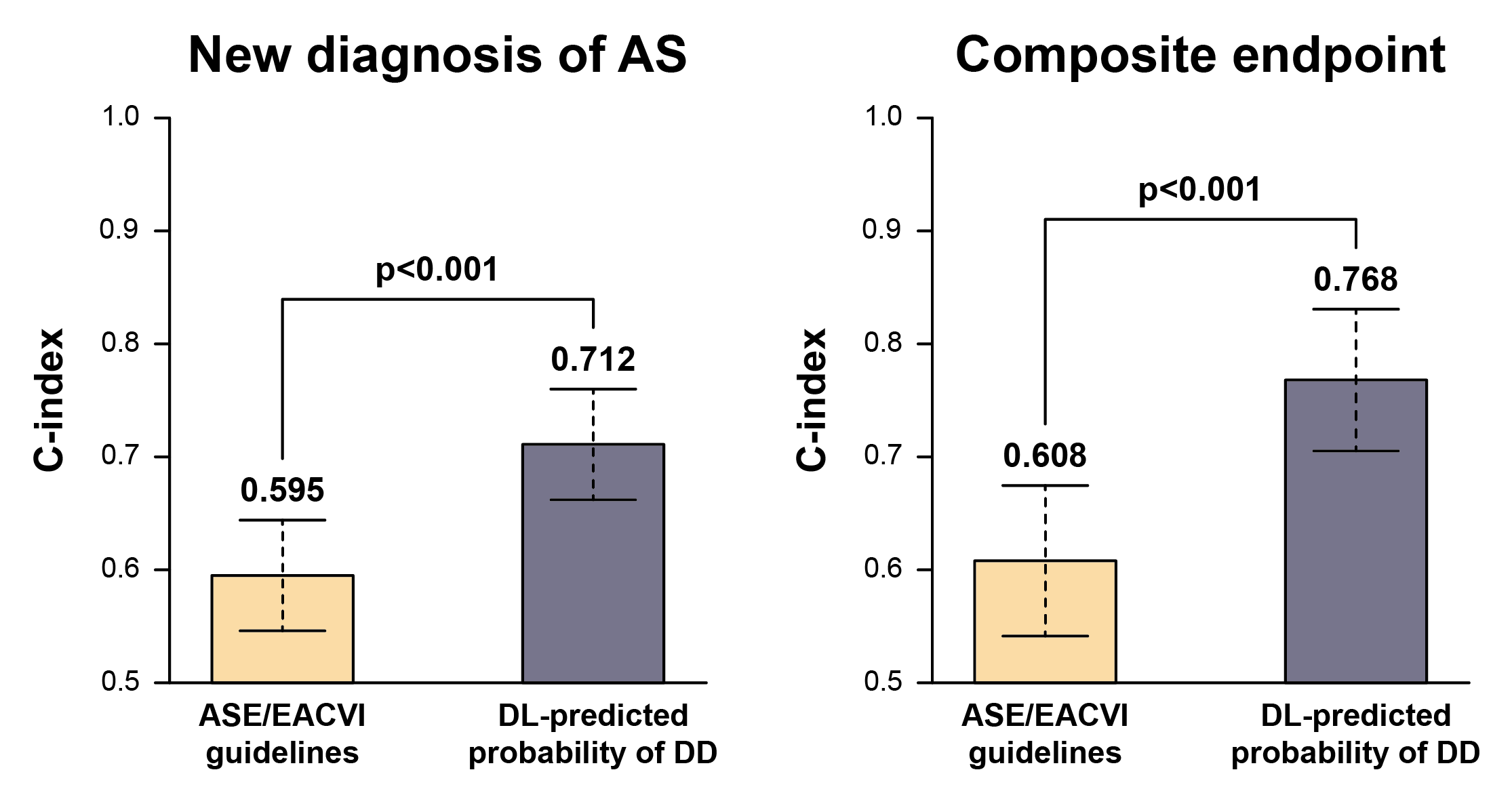

**Supplemental Figure 2** Comparing the prognostic power of the guideline-based DD grading and the DL-predicted probabilities in the ARIC cohort

AS – aortic stenosis, ASE – American Society of Echocardiography, DD – diastolic dysfunction, DL – deep learning, EACVI – European Association of Cardiovascular Imaging
